# Hemodynamics Indicates Differences Between Patients With And Without A Stroke Outcome After Left Ventricular Assist Device Implantation

**DOI:** 10.1101/2023.08.03.23292572

**Authors:** Akshita Sahni, Sreeparna Majee, Jay D. Pal, Erin E. McIntyre, Kelly Cao, Debanjan Mukherjee

## Abstract

Stroke remains a leading cause of complications and mortality in heart failure patients treated with a Left Ventricular Assist Device (LVAD). Hemodynamics plays a central role underlying post-LVAD stroke risk and etiology. Yet, detailed quantitative assessment of hemodynamic variables and their relation to stroke outcomes in patients on LVAD support remains a challenge. Modalities for pre-implantation assessment of post-implantation hemodynamics can help address this challenge. We present an *in silico* hemodynamics analysis for a digital twin cohort 12 patients on LVAD support; 6 with reported stroke outcomes and 6 without. For each patient we created a post-implant twin with the LVAD outflow graft reconstructed from cardiac-gated CT images; and a pre-implant twin of an estimated baseline flow by removing the LVAD outflow graft and driving flow from the aortic valve opening. Hemodynamics was characterized using descriptors for helical flow, vortex generation, and wall shear stress. We observed higher average values for descriptors of positive helical flow, vortex generation, and wall shear stress, across the 6 cases with stroke outcomes when compared with cases without stroke. When the descriptors for LVAD-driven flow were compared against estimated pre-implantation flow, extent of positive helicity was higher, and vorticity and wall shear were lower in cases with stroke compared to those without. Our study suggests that quantitative analysis of hemodynamics after LVAD implantation; and hemodynamic alterations from a pre-implant flow scenario, can potentially reveal hidden information linked to stroke outcomes during LVAD support. This has broad implications on understanding stroke etiology; and using patient digital twins for LVAD treatment planning, surgical optimization, and efficacy assessment.

## 1 Introduction

Heart failure remains a major public health concern, affecting at least 26 million individuals globally [1]. According to the latest American Heart Association reports, over 6 million adults had advanced heart failure between 2007 and 2020 within the United States alone [2]. Left ventricular assist devices (LVADs) have emerged as the standard of care for bridge-to-transplant as well as destination therapy, for patients with advanced heart failure [3–6]. Recent developments in LVAD technology have substantially improved device durability, reduced postoperative adverse events, and improved long-term survival [7]. Despite these advances in LVAD technology, ischemic stroke remains a significant cause of morbidity and mortality [8, 9]. Stroke rates after LVAD implantation have been reported in the range of 11-47% of cases across several studies [10–13]. Although clinical trials with newer generation LVADs have demonstrated reduced stroke rates relative to previous devices [14, 15], a significant number of patients still sustain thrombotic and embolic events leading to stroke. Advancements in understanding the factors that underlie stroke, and reducing the likelihood of post-implant stroke or adverse cerebrovascular events, can therefore substantially impact treatment strategies and improve post-operative quality of life, as also noted in prior works [16, 17]. One major challenge is that the factors that influence the likelihood or etiology of stroke after LVAD implantation are not well understood. Although the mere presence of a metallic foreign body in the blood stream means that a thrombotic risk will always be present (*despite systemic anticoagulation*); yet, some patients will sustain emboli to the brain (*resulting in a stroke*), while others may demonstrate emboli to more distal organs (*where clinical impact is generally less severe*). Furthermore, there may be additional sources of thrombosis over and beyond expectations from device-blood contact interactions, such as thrombogenicity due to flow stasis. Both of these aspects have potential origins in the specific nature of spatiotemporal hemodynamics driven by the LVAD. Hence, it is necessary to characterize how LVAD-driven flow underlies the occurrence of stroke in some patients but not in others, with similar device-related likelihood of thrombosis and embolisms.

As motivated above, hemodynamic features are acknowledged to play a key role underlying complications during LVAD circulatory support [18, 19]. Yet, detailed analysis of 3-dimensional space-time varying flow information as a predictor of post-implantation likelihood or origin of stroke is lacking. Standard-of-care imaging does not currently provide such detailed flow information; thereby requiring alternative quantitative flow analysis techniques to comprehensively characterize blood flow driven by an LVAD. Advances in *in silico* modeling, based on integration of computational hemodynamics modeling with patient-specific anatomy and medical or physiological data, have become a viable avenue for such quantitative analysis; leading to the recent emphasis on Virtual or Digital Twins for personalized models of various healthcare scenarios [20–22]. Several previous studies on circulatory support for LVAD have used computational fluid dynamics to investigate aortic hemodynamics as a function of LVAD outflow graft anastomosis features such as location and angle with the aorta [23–31]. These investigations have specifically looked at factors such as hemodynamic patterns, thrombogenicity, and aortic valve reopening. Through a high-resolution digital representation of the patient with an LVAD, these types of models adopt the concept of a digital twin representation of the post-implant hemodyamic scenarios. The vast majority of these analyses focus on hemodynamics after LVAD implantation, and little is known about: (1) identifying the link between specific flow descriptors and actual stroke/non-stroke outcomes; as well as (2) analysis of (*or comparisons against*) hemodynamic information pre-implantation. Our hypothesis is that the perturbations in flow patterns related to LVAD support can inform the risk of stroke. In a series of prior works [32, 33], we have developed a computational framework to quantitatively assess a range of hemodynamic descriptors such as helicity, vorticity, wall shear, and energy dissipation both as: (a) function of LVAD outflow graft anastomosis; and (b) differences with an estimated pre-implant flow without an operating LVAD. Here we seek to specifically address how the hemodynamics post-implantation, and hemodynamic alterations from a pre-implant flow scenario, are related to stroke risks post-implantation. For this, we present a retrospective secondary analysis on a 12 patient cohort, with 6 having reported stroke outcomes, and 6 without stroke. For each, a patient-specific data-integrated virtual twin model for the pre- and post-implant scenario is reconstructed, to create digital versions of each cohort. Our goal is to quantify detailed differences in hemodynamics between stroke and no-stroke outcomes using commonly used flow descriptors, to elucidate whether hemodynamics holds hidden information pertaining to stroke outcomes in subjects on LVAD circulatory support, regardless of device type.

## 2 Methods

### 2.1 Patient selection

Patient cases were selected from an existing clinical database of patients undergoing LVAD implantation at the University of Colorado Anschutz Medical Campus. For this study, we utilized de-identified patient data from this database for retrospective secondary analysis. Hence, the study was declared to be exempt from IRB review by the Colorado Multiple Institutional Review Board (COMIRB). For each patient, we collected cardiac gated CT angiography scans (*gated to diastole*) with the operating LVAD in place, along with report of clinical findings and key clinical metrics. A total of 12 patient cases were selected for this study. Patient cases were selected based on practical considerations for image-based modeling, such that: (a) CT image quality, ensuring no artefacts from the LVAD; (b) patient case had both full aortic arch upto first level branch vessels clearly visible in the CT scan; (c) and the LVAD outflow graft anastomosis is clearly visible in the CT scan. Key patient clinical variables used for this study have been listed in Table 1. Cases were categorized into two sets with 6 patients each. The first category included 6 patients who experienced a stroke post LVAD treatment (*identified as a cerebrovascular accident or CVA in this study*); while the second category included 6 patients who did not experience any cerebrovascular incidents post LVAD implantation (*identified using the label non-CVA in this study*). Patients reporting a stroke outcome also underwent non-contrast and contrast enhanced head-neck CT imaging. Time between VAD surgery and head-neck imaging was used as an estimate for time between surgery and stroke (listed in Table 1). Furthermore, as per clinical records, these patients had no reported levels of aortic valve opening, hence we assumed that the aortic valve was sealed close throughout this study, an assumption that we have discussed at length in our prior works [32, 33].

**Table 1:** A listing of relevant clinical variables for the 12 patient cases included in this study. Gender is indicated as Male (M) or Female (F). The various VAD types (†) are: HM 2 - HeartMate II; HM 3 - HeartMate III; HW - HeartWare. VAD RPM was noted at the time of chest imaging using cardiac gated CT. The Cases listed here, and subsequent tables and figures are used consistently to identify and group patients with respect to stroke and non-stroke outcomes. Time to CVA is estimated in days between LVAD surgery and the first post-stroke head-neck imaging. For case C1; patient needed a pump replacement, and time to CVA was estimated from final pump placement date.

| Case | C1 | C2 | C3 | C4 | C5 | C6 | N1 | N2 | N3 | N4 | N5 | N6 |
| --- | --- | --- | --- | --- | --- | --- | --- | --- | --- | --- | --- | --- |
| Gender | M | M | F | M | M | M | M | M | F | F | M | M |
| Age at implant | 46-50 | 61-65 | 56-60 | 66-70 | 36-40 | 56-60 | 61-65 | 61-65 | 61-65 | 61-65 | 56-60 | 71-75 |
| VAD type<sup>†</sup> | HM 2 | HW | HM 2 | HM 3 | HW | HW | HM 3 | HW | HM 3 | HM 3 | HW | HM 3 |
| VAD RPM | 9400 | 2500 | 9000 | 4800 | 2560 | 2720 | 5700 | 2720 | 5500 | 5600 | 2700 | 5400 |
| MAP (mmHg) | 84 | 82 | 66 | 66 | 88 | 90 | 85 | 73 | 92 | 80 | 78 | 78.5 |
| VAD Flow (l/min) | 6.9 | 4.7 | 4.1 | 3.8 | 3.7 | 3.7 | 5.3 | 5.3 | 3.7 | 4.0 | 3.6 | 4.0 |
| Stroke (CVA) | Yes | Yes | Yes | Yes | Yes | Yes | No | No | No | No | No | No |
| Time to CVA (days) | 553* | 442 | 69 | 1 | 34 | 18 | — | — | — | — | — | — |

### 2.2 Image-based modeling of patient vasculature

The cardiac gated CT angiography scans were used to generate patient-specific 3D vascular models using the open source software tool SimVascular [34]. Specifically, from each patient CT image stack, pathlines were created for the aorta, first generation branch vessels, and the LVAD outflow graft. Thereafter, a 3D model of the vascular region of interest, with the LVAD outflow graft attached in place, was generated by creating planar 2D segmentations of the vessel and LVAD outflow graft lumen, and lofting the segmentations. The workflow for generating the 3D vascular model from CT images is presented for one of the cases (Case C6) in Figure 3. Illustration of all the 12 patient anatomy models is presented in Figure 4, with their de-identified case numbers referring to the original anonymized record IDs used in the study, and the models categorized as *CVA* and *non-CVA*. To ensure that the LVAD outflow graft is reconstructed in a geometrically consistent manner, graft inner diameter was measured at different locations along the graft centerline, and the resulting diameters were verified against known standard values as per manufacturer specifications for the device. The comparison of these diameters is shown in Figure 5. This comparison was done to ensure that the LVAD outflow graft for each case is reconstructed in a geometrically consistent manner. An additional set of 12 models were generated by eliminating the path and segmentations for the LVAD outflow graft for each patient, and re-lofting the arterial network without the LVAD graft. These models were referred to as the *Baseline Model* for each patient case, representative of the vascular geometry before LVAD implantation (*See Figure 3* *for schematic illustration*). As established in our prior works [32], the baseline cases were included to generate estimates of arterial hemodynamics prior to LVAD implantation, and compare this pre-implantation flow estimate with post-implantation flow to quantify the hemodynamic alterations experienced due to LVAD operation. The vessel lumen for each of these 24 total models were discretized into a computational grid comprising linear, tetrahedral element, and the maximum edge sizes for each model are listed in Table 2. The edge sizes are informed by mesh refinement and aortic hemodynamic studies reported in several prior works [35, 36].

**Table 2:** A description of all derived hemodynamic parameters that are used for the purpose of specifying the computational hemodynamics simulations for each of the 12 cases studied here. Part a. presents the Total Arterial Resistance (TAR). Part b. represents all the branching artery resistance boundary condition values. Methodology for both are provided in Section 2. Part c. identifies the computational grid/mesh size and time-step size for each model.

| Case | C1 | C2 | C3 | C4 | C5 | C6 | N1 | N2 | N3 | N4 | N5 | N6 |
| --- | --- | --- | --- | --- | --- | --- | --- | --- | --- | --- | --- | --- |
| Stroke (CVA) | Yes | Yes | Yes | Yes | Yes | Yes | No | No | No | No | No | No |
| **Part a.** | **Derived total arterial resistance (TAR) (g.mm<sup>-4</sup>.sec<sup>-1</sup>)** |  |  |  |  |  |  |  |  |  |  |  |
| Resistance (TAR) | 0.10 | 0.14 | 0.13 | 0.14 | 0.19 | 0.19 | 0.13 | 0.11 | 0.20 | 0.16 | 0.17 | 0.15 |
| **Part b.** | **Resistance from branching artery boundary condition (g.mm<sup>-4</sup>.sec<sup>-1</sup>)** |  |  |  |  |  |  |  |  |  |  |  |
| R. Subclavian | 0.862 | 1.163 | 2.144 | 1.049 | 1.283 | 1.772 | 1.568 | 0.876 | 4.690 | 1.839 | 0.739 | 1.008 |
| L. Subclavian | 0.810 | 1.577 | 2.614 | 2.654 | 1.339 | 3.842 | 2.045 | 1.280 | 6.572 | 2.629 | 1.076 | 1.483 |
| R. Carotid | – | 1.981 | 1.899 | 2.351 | – | 2.123 | 2.470 | 1.369 | 3.853 | 1.154 | – | 3.843 |
| L. Carotid | 0.978 | 2.126 | 2.468 | 2.453 | 1.334 | 3.257 | 3.680 | 1.630 | 3.734 | 1.758 | 2.948 | 3.139 |
| Desc. Aorta | – | – | 0.274 | 0.305 | 0.336 | – | 0.258 | – | 0.242 | – | 0.469 | 0.359 |
| Celiac Hepatic | 1.762 | 2.219 | 2.232 | 3.215 | – | 8.864 | 6.775 | 3.897 | – | 5.189 | – | 5.665 |
| Splenic | 1.621 | 1.607 | 4.007 | 1.559 | – | 3.467 | 1.460 | 1.157 | – | 2.029 | – | 2.936 |
| Sup. Mesenteric | 1.257 | 1.179 | 1.590 | 1.247 | – | 3.051 | 2.159 | 1.421 | – | 2.921 | 1.672 | 2.259 |
| R. Renal | 2.420 | 5.076 | 2.169 | – | – | 5.939 | 2.491 | 1.646 | – | 4.756 | 7.470 | 6.920 |
| L. Renal | 1.859 | 3.441 | 1.816 | – | – | 2.262 | 2.902 | 1.286 | – | 2.716 | 3.543 | 2.543 |
| R.Ext. Fem. | 0.784 | 0.804 | – | – | – | 1.164 | – | 0.826 | – | 1.656 | – | – |
| L.Ext. Fem. | 0.703 | 0.958 | – | – | – | 1.154 | – | 0.720 | – | 1.827 | – | – |
| R. Profunda | 1.837 | – | – | – | – | 4.456 | – | – | – | 1.622 | – | – |
| L. Profunda | 1.472 | – | – | – | – | 4.050 | – | – | – | 1.936 | – | – |
| **Part c.** | **Spatial and temporal discretization parameters** |  |  |  |  |  |  |  |  |  |  |  |
| Max. edge size (mm) | 0.671 | 0.775 | 0.873 | 0.671 | 0.715 | 0.785 | 0.752 | 0.814 | 0.794 | 0.897 | 0.837 | 0.850 |
| Num. elements (millions) | 15.15 | 7.60 | 5.91 | 12.51 | 4.05 | 11.26 | 11.04 | 7.80 | 5.23 | 7.30 | 7.12 | 9.33 |
| Time step size (sec) | 0.001 | 0.001 | 0.001 | 0.001 | 0.001 | 0.001 | 0.001 | 0.001 | 0.001 | 0.001 | 0.001 | 0.001 |

**Figure 1.**
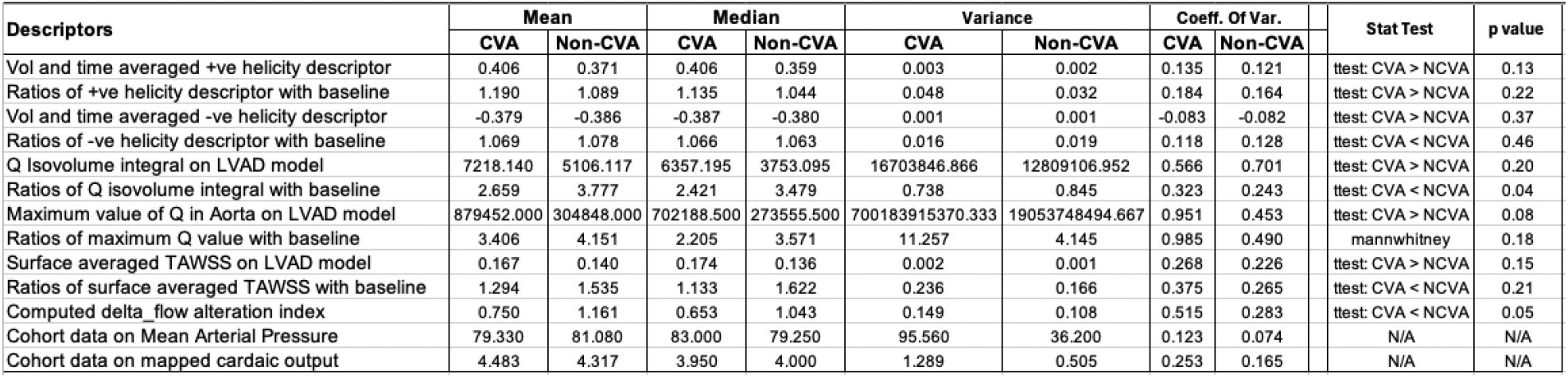
Compilation of individual descriptor data and p-values for the various statistical comparisons as conducted in this study.

**Figure 2.**
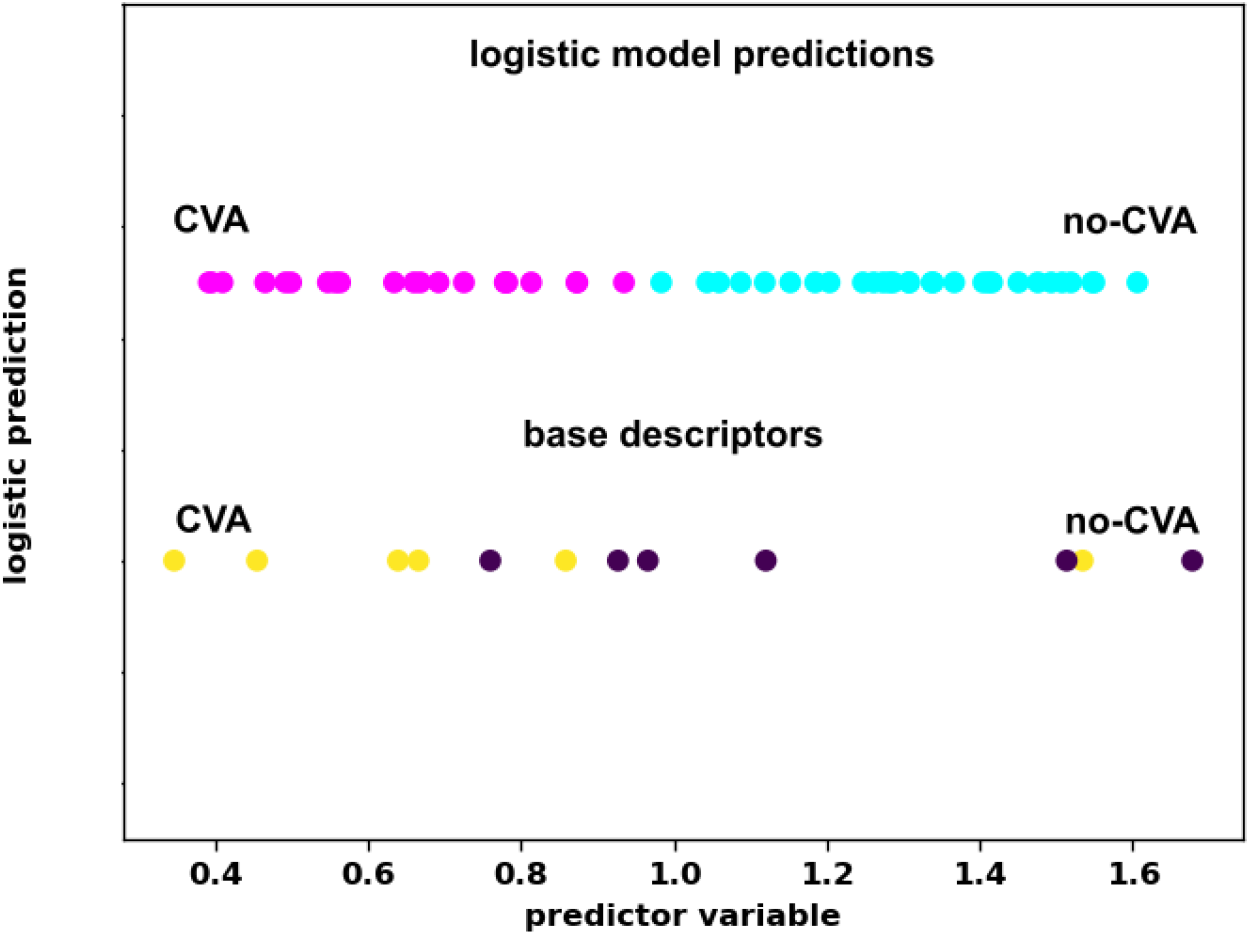
Example of a sample logistic regression modeling with stroke outcome as a categorical variable, and the hemodynamic alteration variable with baseline Δ_flow_ as a predictor/independent descriptor variable.

**Figure 3.**
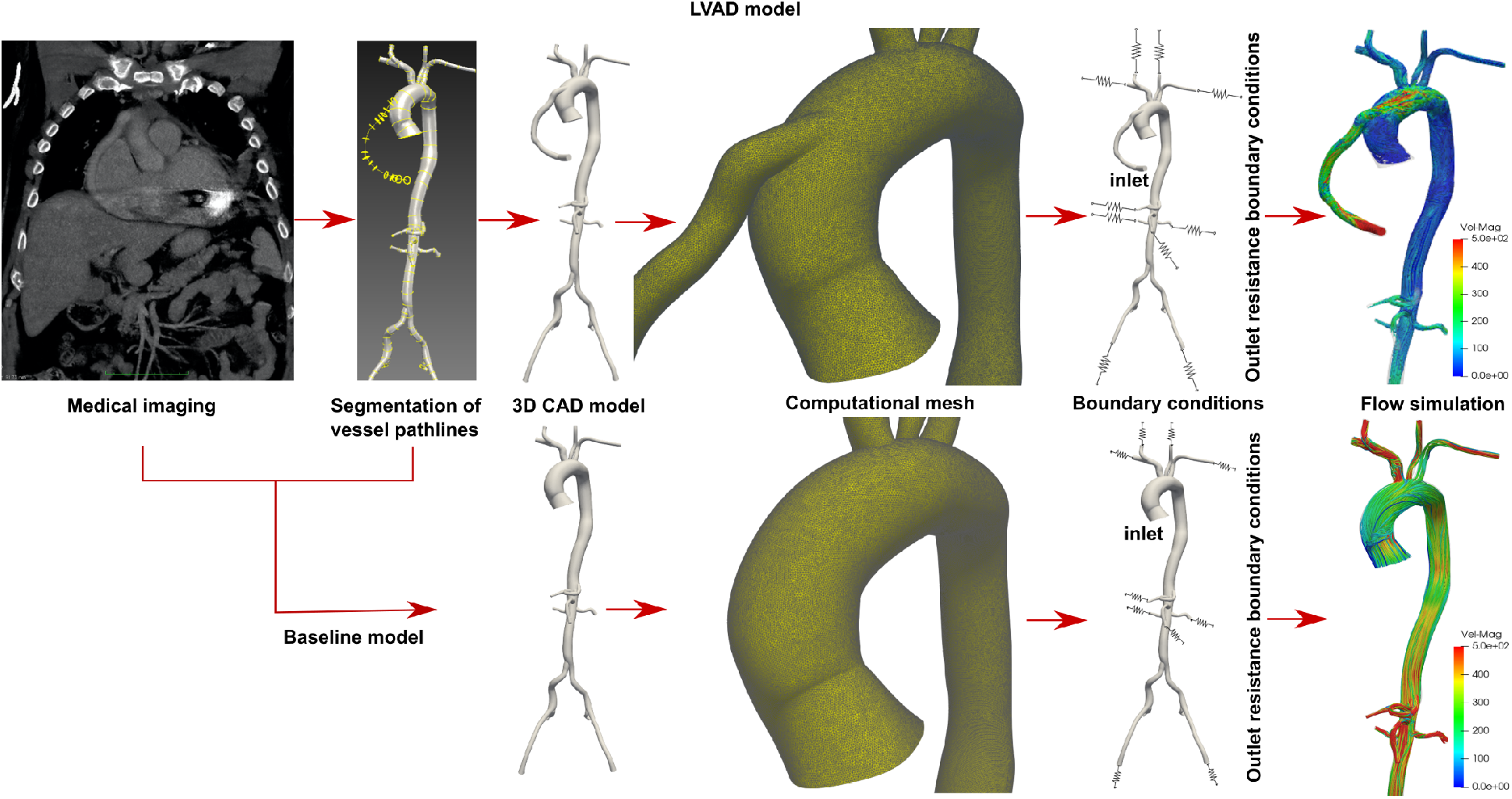
Schematic overview of the sequence of steps in creating 3D patient vascular models with resistance based outflow boundary conditions from CT image with LVAD graft and baseline model.

**Figure 4.**
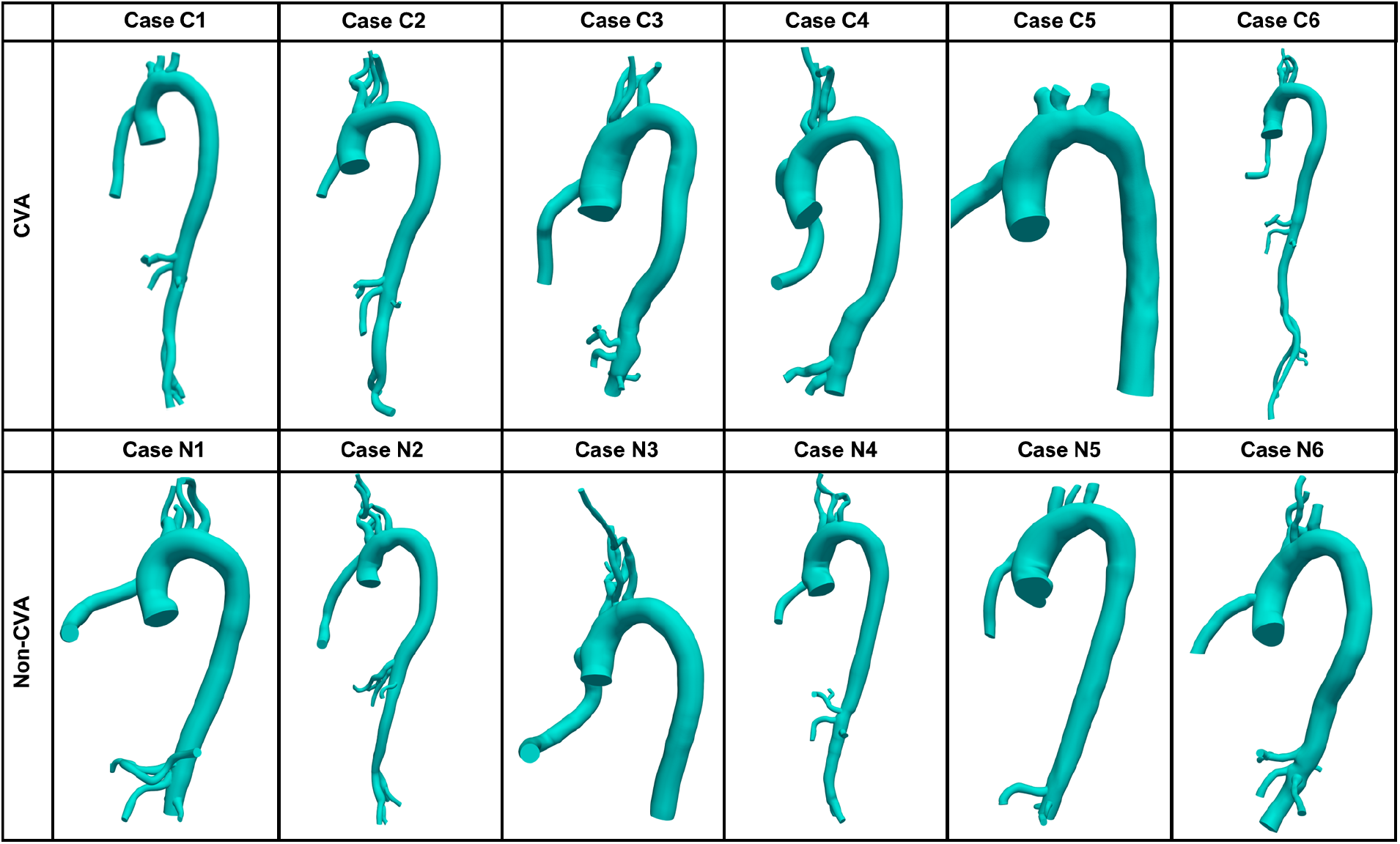
An illustration of all 12 patient vascular models as reconstructed from CT images, with the LVAD graft of the operational LVAD attached. The Case IDs listed here are the same as identified in Table 1.

**Figure 5.**
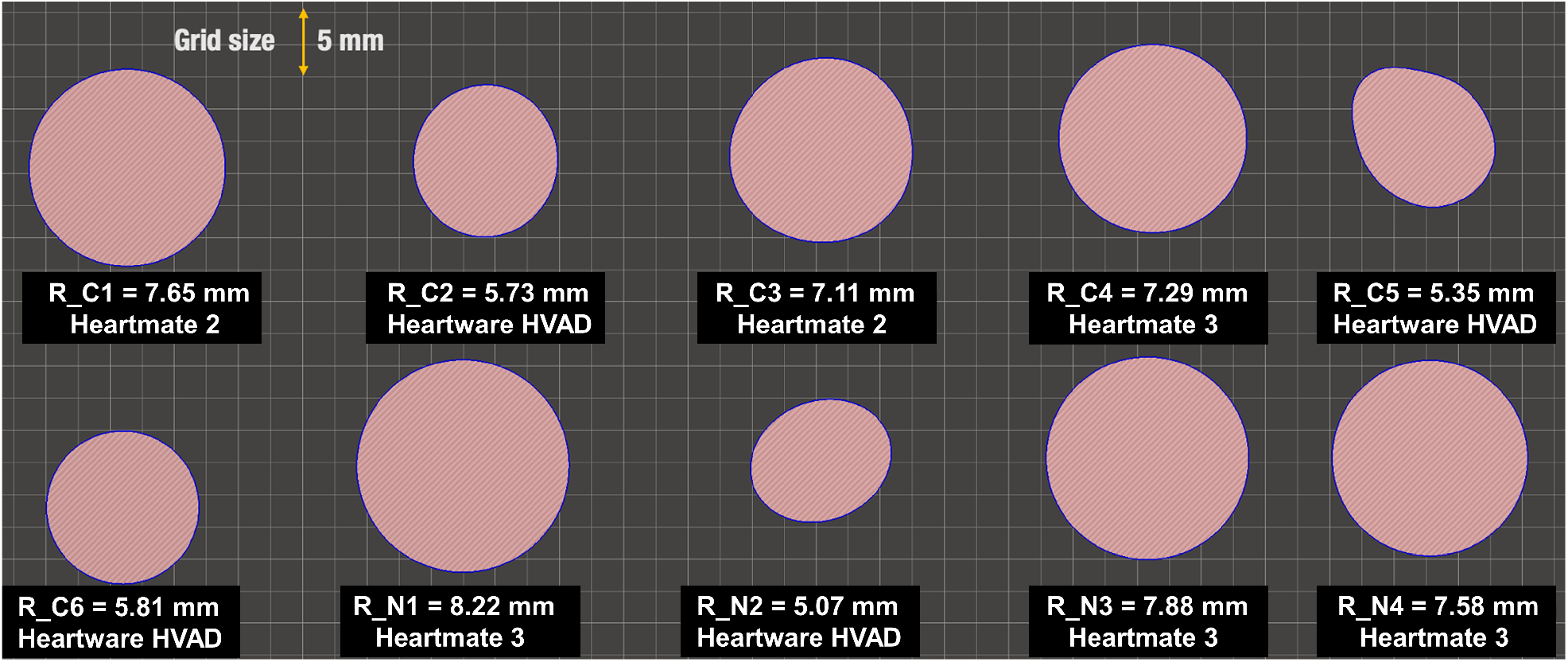
LVAD outflow graft diameters measured from the twelve reconstructed LVAD patient models, illustrated here for verification of graft reconstruction. We note that Heartmate 2/3 grafts are known to be (approx) 10 mm in diameter, while Heartware HVAD grafts are known to be (approx) 14 mm in diameter. Our reconstructed grafts are in the range of 2-16% different from the 10/14 mm sizes respectively as noted above. This indicates good reconstruction quality, since these grafts are known to also expand upto 10% due to flow pressure. See also Supplementary Material.

### 2.3 Patient-specific hemodynamics modeling

Blood was assumed to be a Newtonian fluid demonstrating a linear relationship between shear stress and strain rate [37], with a constant effective average density of 1.060× 10^−3^ g.mm^3^ and effective dynamic viscosity 0.004 g.mm^-1^.sec^-1^. Vessel walls were assumed rigid, and for the LVAD models the aortic valve was assumed to be closed as stated in Section 2.1. Blood flow through all models was computed by solving the Navier-Stokes equations and the continuity equation for fluid momentum and mass balance respectively, using a Petrov-Galerkin stabilized linear finite element method [38–41]. The computations resulted in spatiotemporally varying velocity and pressure fields in the patient’s arterial pathway. For each patient the LVAD outflow graft was assumed to drive uniform continuous flow, mapped onto a parabolic profile across the outflow graft inlet, without any pulse modulation. The VAD flow rate (*Q*_*V* *AD*_) and Mean Arterial Pressure (*MAP*) was obtained from patient records (see Table 1), and the Total Arterial Resistance (*TAR*) was estimated as follows (*see Table 2* *for estimated values for each patient*):

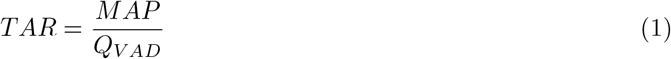

Across all *CVA* cases the average MAP was 79.33 mmHg and average mapped cardiac output was 4.48 l/min. Likewise, across all *non-CVA* cases the average MAP was 81.08 mmHg and average mapped cardiac output was 4.32 l/min. Following this, we assumed that flow into each major branching vessel was approximately proportional to the cross-sectional areas of the vessel outlets, based on the assumption of minimal work [42]. Using this estimated flow distribution targets, the TAR was decomposed to assign individual outlet resistance values to the outlets for each patient model, as illustrated in Figure 3. For the baseline models, the *Q*_*V* *AD*_ was instead imposed as the cardiac output exiting from the aortic valve into the aorta of the model. The *Q*_*V* *AD*_ was mapped onto the aortic inlet face as a parabolic profile function in space; and mapped across time using a previously used physiologic pulse-profile shape [32] (*thereby scaling the profile to match the Cardiac Output to be equal to Q*_*V* *AD*_ *for each subject*). The target flow distribution and vessel outlet resistances to all other outlet boundary faces were kept the same across the LVAD and baseline models. Table 2 summarizes the resistance values for the first generation arteries for each patient case. Using these boundary conditions, blood flow through each of the 12 patient models with VAD, and the respective baseline was simulated for a total duration of 3 seconds, with a numerical integration time-step of 0.001 seconds. Once completed, simulated results for the third second of the total simulation duration were extracted and further post-processed to obtain relevant hemodynamic descriptors.

### 2.4 Hemodynamic descriptors

For quantitative analysis of the hemodynamics across the 12 LVAD and baseline models, we used Eulerian flow descriptors based on three widely adopted hemodynamic features: helicity, wall shear, and vorticity. These descriptors take micron-scale space-time varying flow feature information, and coalesce them into a single aggregate macroscale parameter that averages over the representative region of interest. Detailed description, rationale, and definitions have been outlined in our prior work [32]. Here, we briefly reproduce key details and definitions for describing our analysis. The first descriptor characterizes helical flow patterns generated due to aortic curvature and tortuosity. We computed a time-averaged localized normalized helicity 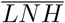 defined as follows [32, 43]:

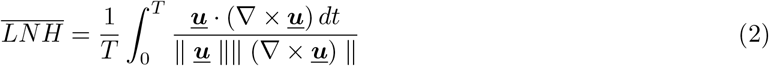

where ***u*** is the computed blood flow velocity. Based on the above definition, we further devised a volume integrated helicity metric Φ_*H*_ and volume averaged helicity metric ⟨Φ_*H*_ ⟩ defined as follows for a given range of averaged 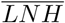 values [*L*_1_, *L*_2_]:

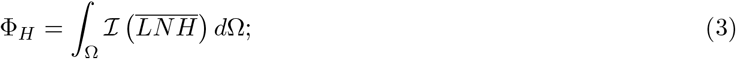

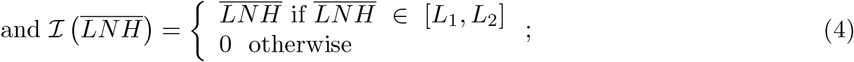

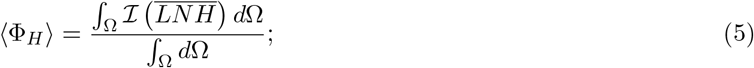

where the volume averaging operation was conducted on the aortic arch alone, excluding the LVAD outflow graft and the first generation carotid and subclavian branches. As a second hemodynamic descriptor, we used the Q-criteria to characterize extent of vortical structures developed in the aortic arch caused by flow emanating from the LVAD outflow graft. *Q*-criteria is a quantitative descriptor that mathematically identifies locations where rotational flow dominates straining flow [44]. We computed the Q-criterion value based on computed velocity field ***<u>u</u>*** as:

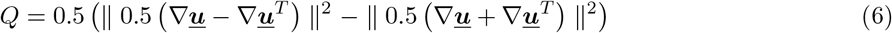

For our analysis, the above definition was used to compute a volume averaged Q-criterion metric ⟨Φ_*Q*_ ⟩ defined as follows [32]:

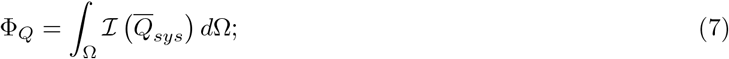

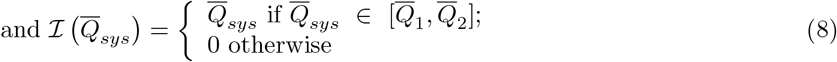

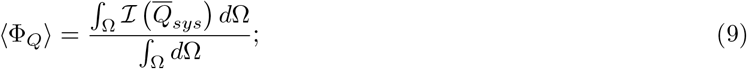

where Ω denotes the integration domain, which is the aortic arch region excluding other branches; 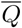 denotes the *Q*-value normalized against the systolic *Q*-value for the baseline model (at *t* = 0.18*s* of the third cycle of simulated flow). Lastly, we compared the extent of wall shear stresses along the aortic wall by computing the time averaged state of wall shear stress (TAWSS) from the computed flow velocity field ***u*** as follows:

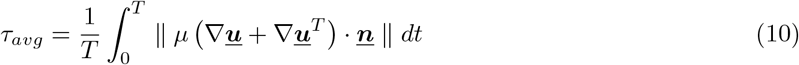

where ***<u>n</u>*** denotes the normal vector along the wall of the aorta, and *T* = 1.0*s* or one cycle for time averaging. Based on the above definition, a surface averaged TAWSS descriptor was computed as follows:

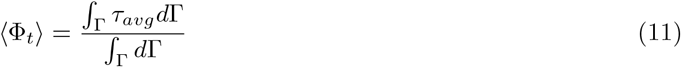

where Γ denotes the surface of the aortic arch lumen, excluding any branching vessels. Readers are referred to our prior work [32, 33] for additional details on each of these hemodynamic descriptors. The descriptors were computed using custom post-processing computer programs written in Python using the Visualization Toolkit (VTK) library, and visualized in Paraview.

## 3 Results

### 3.1 Visualization of aortic hemodynamic patterns

Figure 6 illustrates the simulated hemodynamic patterns for the 12 cases with an operating LVAD considered in this study, visualized using streamtubes generated at the *t* = 0.18 second time-point of the third simulation cycle (*corresponding to the timepoint of the peak systole in the third cardiac cycle for the pulsatile baseline flow*). The top row of Figure 6 depicts all *CVA* cases, and the bottom row depicts all *non-CVA* cases. With a non-functional aortic valve, aortic hemodynamics is driven by the impingement of the LVAD outflow jet on the aortic wall. At the jet impingement point there is a spike in pressure associated with local stagnation of flow, and post-impingement the flow is highly non-linear with significant vortex generation and transport into the arch and descending aorta. As this flow progresses beyond the impingement point, the curvature and tortuosity of the aorta also generates secondary circulatory flow which ultimately leads to a state of helical flow, as indicated by the coiling of streamtubes in the aorta in Figure 6. These flow features can be observed over time in the animations provided in the supplementary dataset provided alongwith this article. Additionally, the presence of a non-functional aortic valve causes flow stagnation at the root of the proximal aortic arch, indicated by the prominent slow or arrested flow zones at the aortic root in Figure 6. The corresponding visualization of the estimated pre-implant or *Baseline* hemodynamics for the 12 models is provided in the supplementary dataset.

**Figure 6.**
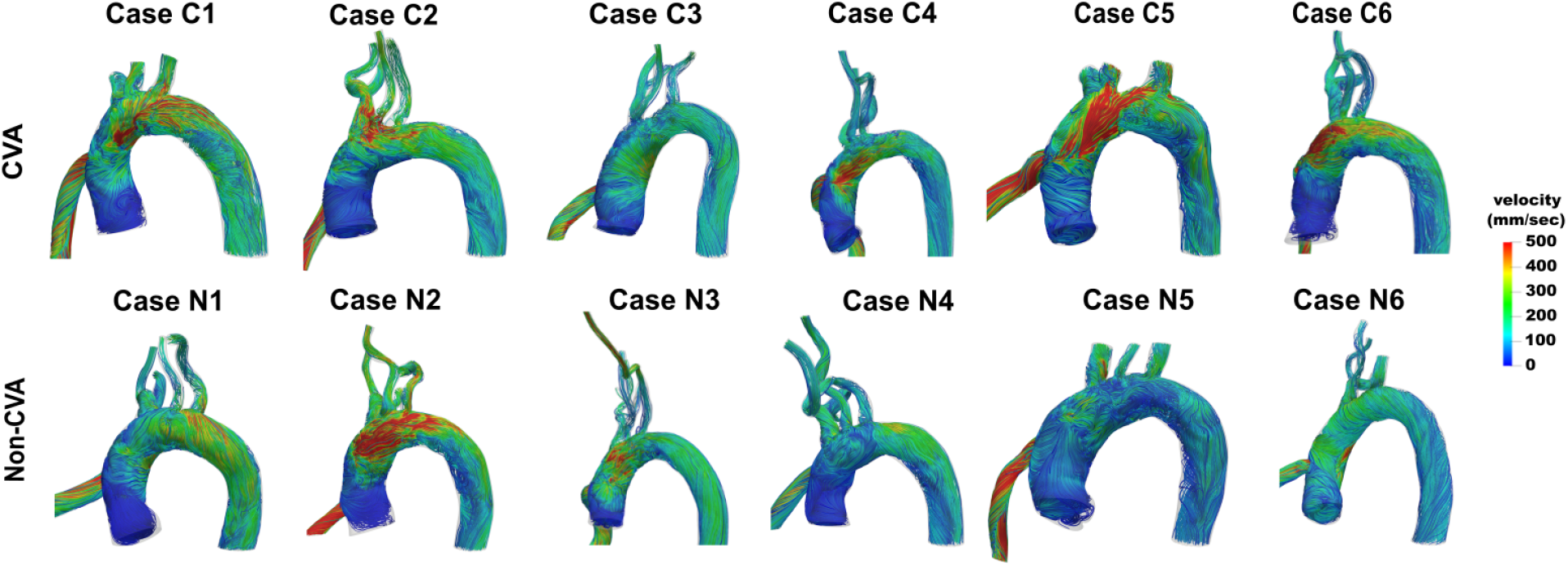
Visualization of spatially varying aortic hemodynamic patterns using velocity streamtubes illustrated for stroke (CVA) cases at the top and non-stroke (non-CVA) cases at bottom. Streamtubes are generated at t = 0.18s within the third simulation cycle, which maps to the timepoint of the peak systole in the third cardiac cycle for the corresponding pulsatile baseline flow.

### 3.2 Comparison of helical hemodynamics in the aorta

Analysis and comparison of helical hemodynamics in the aorta was undertaken for the *CVA* cases and the *non-CVA* cases using the descriptors outlined in Section 2.4. Figure 7.a. depicts the computed averaged helicity descriptor ⟨Φ_*H*_⟩ defined in Section 2.4, estimated for positive helicity ranges [*L*_1_, *L*_2_] = [0.2, 1.0], compared between the *CVA* and the *non-CVA* cases considered in this study. The results show a higher mean value for ⟨Φ_*H*_⟩ for *CVA* cases (*mean=0*.*41*) compared to *non-CVA* cases (*mean=0*.*37*). This indicates that aortic hemodynamics driven by LVAD is potentially associated with greater extent of positively helical flow for cases with a stroke outcome, compared to cases without a stroke outcome. Figure 7.b. depicts the ratio of the computed helicity descriptor ⟨Φ_*H*_⟩ between all models with an operating LVAD and the respective pre-implantation *baseline* models. As observed in our prior work [32], we find that hemodynamics in all models with an operating LVAD has increased positive helicity when compared to the estimated baseline flow (*i*.*e*. ⟨Φ_*H*_ ⟩ *ratios for LVAD vs baseline greater than 1*.*0*). Additionally, for the cases considered here, the ratio of ⟨Φ_*H*_⟩ with baseline flow had a higher mean value for cases with a stroke (*mean = 1*.*19*) compared to cases without a stroke (*mean = 1*.*09*). Figure 7.c. depicts the isosurfaces of LNH values taken at +0.2 and -0.2 for each of the 12 LVAD models. These specific values were chosen to effectively capture helical features that persist even when helicity is averaged over time, as also noted in our prior work [32]. This illustration qualitatively indicates that the extent of helicity varies with patient aorta geometry, and is strongly influenced by the outflow graft anastomosis, as was also observed in our multiparametric study discussed in [32]. The corresponding helicity descriptor values for negative helical flow were also computed using the definitions stated in Section 2.4, with [*L*_1_, *L*_2_] = [−1.0, −0.2], and the comparison is presented in Figure 8. However, unlike positive helicity, we did not observe consistent trends or differences between the *CVA* and the *non-CVA* cases for negative helicity. Table 3 presents a list of descriptive statistics for these quantities for further quantitative reference supporting our observations. Additional details on statistical analysis of the descriptors are included in the appendix to this manuscript.

**Table 3:** A description of summary statistics: mean, median, variance, and coefficients of variation (represented by the ratio between standard deviation and mean) for all the hemodynamic descriptors described in this study. Text in Sections 3.1-3.5 refer to the means, but as observed here, the medians follow the same comparative trends as the means.

| Descriptors | Mean |  | Median |  | Variance |  | Coeff. of var. |  |
| --- | --- | --- | --- | --- | --- | --- | --- | --- |
|  | CVA | Non-CVA | CVA | Non-CVA | CVA | Non-CVA | CVA | Non-CVA |
| Averaged +ve helicity | 0.406 | 0.371 | 0.406 | 0.359 | 0.003 | 0.002 | 0.135 | 0.121 |
| Ratios of +ve helicity w. baseline | 1.190 | 1.089 | 1.135 | 1.044 | 0.048 | 0.032 | 0.184 | 0.164 |
| Averaged -ve helicity | -0.379 | -0.386 | -0.387 | -0.380 | 0.001 | 0.001 | -0.083 | -0.082 |
| Ratios of -ve helicity w. baseline | 1.069 | 1.078 | 1.066 | 1.063 | 0.016 | 0.019 | 0.118 | 0.128 |
| Q Isovolum integral | 7218.140 | 5106.117 | 6357.195 | 3753.095 | 16703846.866 | 12809106.952 | 0.566 | 0.701 |
| Ratios of Q integral w. baseline | 2.659 | 3.777 | 2.421 | 3.479 | 0.738 | 0.845 | 0.323 | 0.243 |
| Maximum Q in Aorta | 879452.000 | 304848.000 | 702188.500 | 273555.500 | 700183915370.333 | 19053748494.667 | 0.951 | 0.453 |
| Ratios of maximum Q w. baseline | 3.406 | 4.151 | 2.205 | 3.571 | 11.257 | 4.145 | 0.985 | 0.490 |
| Surface averaged TAWSS | 0.167 | 0.140 | 0.174 | 0.136 | 0.002 | 0.001 | 0.268 | 0.226 |
| Ratios of averaged TAWSS w. baseline | 1.294 | 1.535 | 1.133 | 1.622 | 0.236 | 0.166 | 0.375 | 0.265 |
| Combined flow alteration $\Delta_{flow}$ | 0.750 | 1.161 | 0.653 | 1.043 | 0.149 | 0.108 | 0.515 | 0.283 |

**Figure 7.**
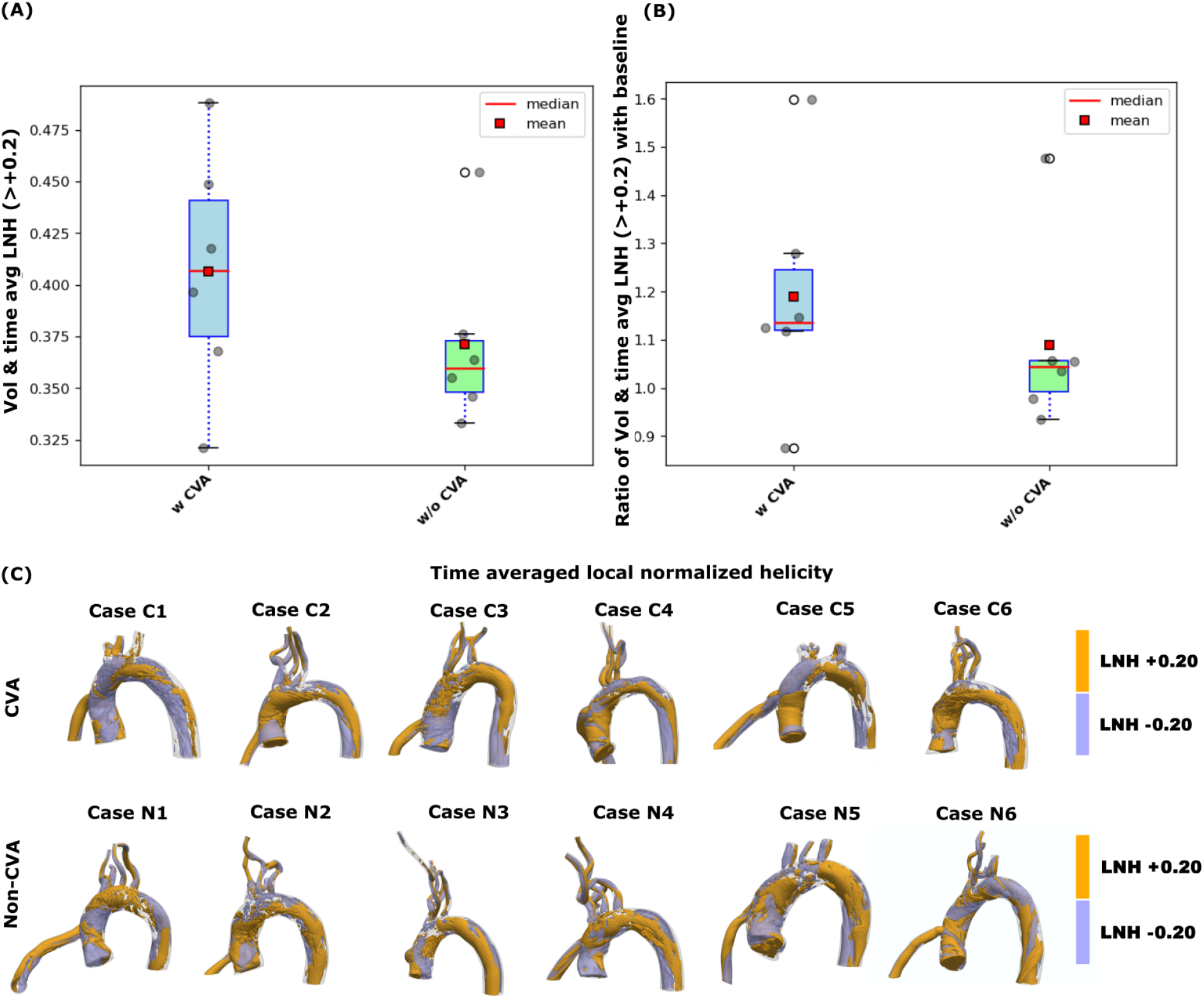
Comparison between trends in time averaged positive local normalised helicity for CVA vs. non-CVA cases: (A) depicts the extent of helicity for flow with LVAD when computed over the arch volume and across one cardiac cycle; (B) depicts the ratio of positive helicity descriptor with that for the baseline model; and (C) presents isosurface visualisations of time averaged local normalised helicity for CVA and non-CVA cases viewed at ±0.20.

**Figure 8.**
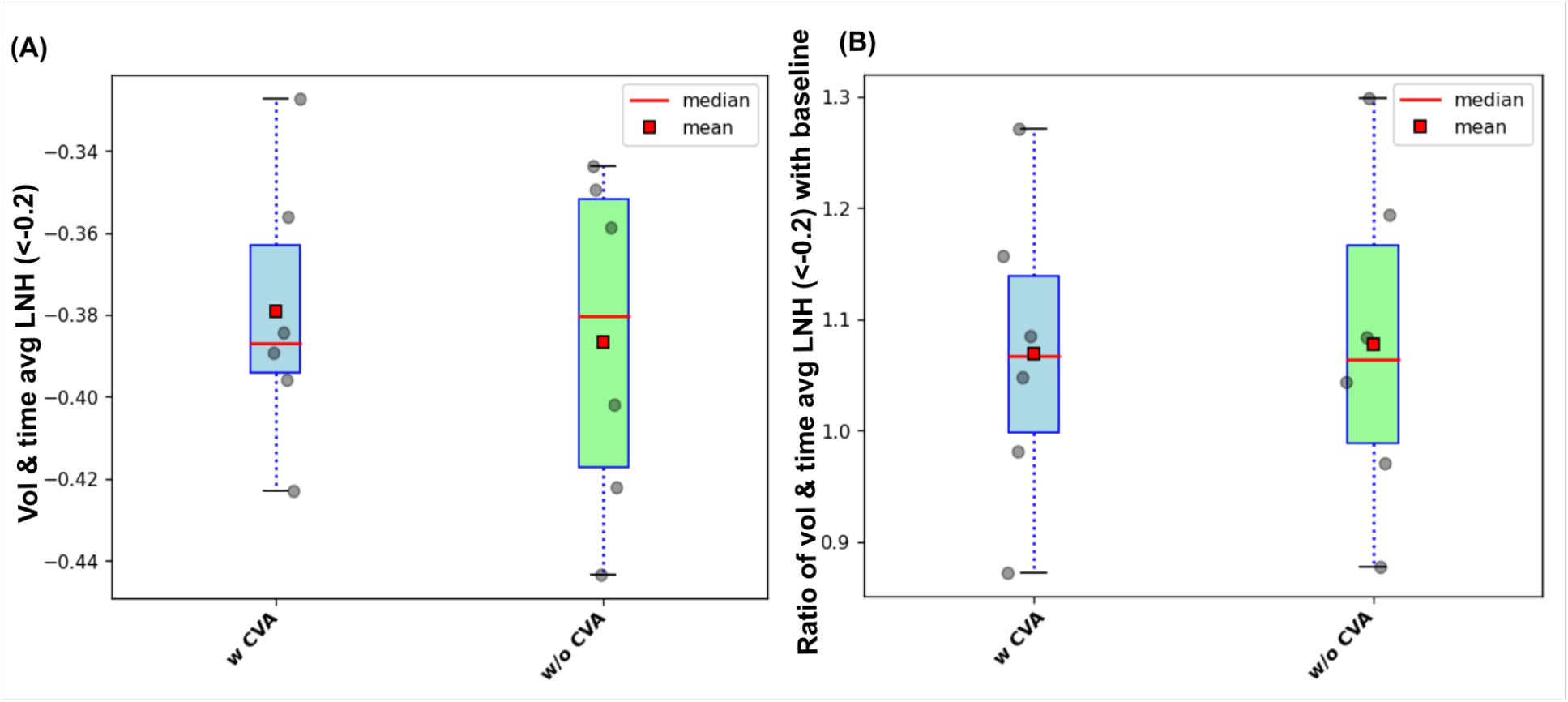
Comparison between trends in time averaged negative local normalised helicity (LNH) results for CVA vs. non-CVA cases: (A) with LVAD over arch volume and one cardiac cycle, (B) Ratio of negative LNH values with baseline model.

### 3.3 Comparison of vortex structures due to jet impingement

Figure 9 illustrates the comparison of vortical flow driven by the LVAD outflow jet between the *CVA* and *non-CVA* cases, using the averaged vorticity descriptor ⟨Φ_*Q*_⟩ defined in section 2.4, as well as the respective maximum Q-criterion values obtained at *t* = 2.18 seconds in the 3 second simulation window (*corresponding to the timepoint of the peak systole in the third cardiac cycle for the pulsatile baseline flow*). For this analysis, 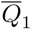 was set at 0.001 and 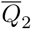 was set at the maximum 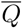 ratio with value 1. Figure 9.a. presents the comparison of ⟨Φ_*Q*_⟩ between the *CVA* and the *non-CVA* models; while Figure 9.c. presents the comparison for the maximum Q-criteria values. For both maximum Q and ⟨Φ_*Q*_⟩, we observe a higher mean for the *CVA* models (*mean* ⟨Φ_*Q*_⟩ = 7218.14; *mean Q*_*max*_ = 879452.000) when compared against the *non-CVA* models (*mean* ⟨Φ_*Q*_⟩ = 5106.12; *mean Q*_*max*_ = 304848.000). This indicates that aortic hemodynamics driven by LVAD is potentially associated with greater extent of vortex dynamics for cases with a stroke outcome, compared to cases without a stroke outcome. Ratio of the averaged vorticity descriptor ⟨Φ_*Q*_⟩and the maximum Q-criteria value with respect to the pre-implantation *baseline* flow estimate is presented in Figure 9.b. and Figure 9.d. Across all 12 models with an attached LVAD outflow graft, both vorticity descriptors were significantly higher than the corresponding baseline models (*all ratios with baseline >* 1.0). This indicates that the extent of vorticity generated due to LVAD outflow jet impingement on the aortic wall is greater than the extent of vorticity in the aortic arch originating from the aortic jet emanating from aortic valve opening. This trend is also consistent with observations generated in our prior parametric study [32]. Additionally, the mean values for both of these ratios were lower for the *CVA* (*mean ratio* ⟨Φ_*Q*_⟩ = 2.66; *mean ratio Q*_*max*_ = 3.41) cases when compared against *non-CVA* cases (*mean ratio* ⟨Φ_*Q*_⟩ = 3.78; *mean ratio Q*_*max*_ = 4.15). This suggests that the baseline flow dynamics with normal aortic inlet may potentially have greater extent of vorticity for cases with a stroke outcome when compared to those without a stroke outcome after LVAD implantation. Figure 9.e. illustrates the instantaneous 3-dimensional spatially varying vortical structures visualized as iso-surfaces of the computed Q-criteria taken at 0.1%*Q*_*max*_ for all the 12 models with an operating LVAD. Qualitative variations in the vortex structures observed in Figure 9.e. can be attributed to differences in jet impingement owing to varying LVAD outflow graft anastomosis angles and varying operating VAD flow rates. Additionally, the regions at the root of the arch void of any vortex structures corresponds to regions of flow stasis as also identified in Figure 6. A compilation of descriptive statistics for these quantities is included in Table 3, and discussion on additional statistical analysis is provided in the appendix to this manuscript.

**Figure 9.**
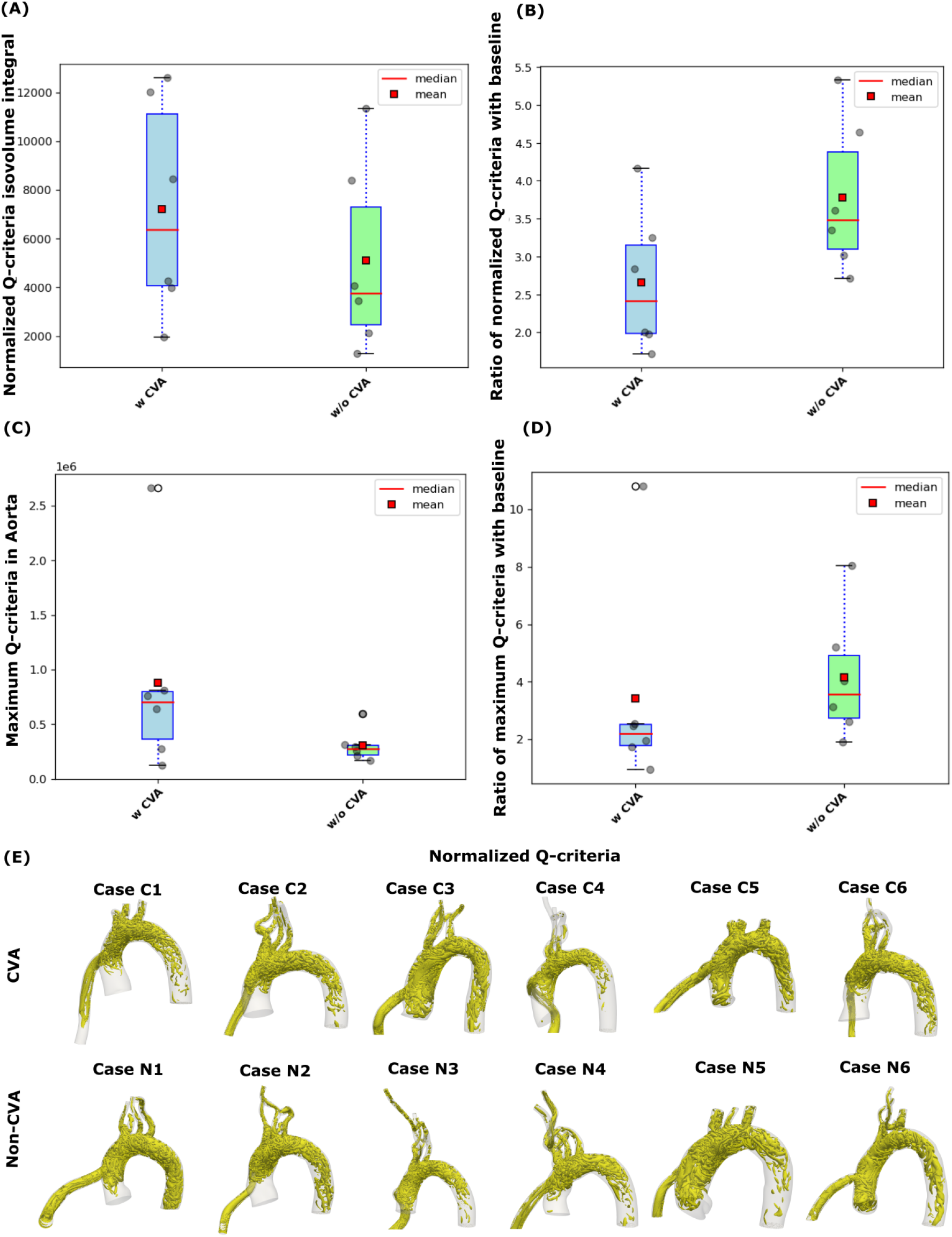
Comparison between trends in Q-criterion for patients with a stroke(CVA) vs. non-stroke(non-CVA) LVAD outcome: (A) depicts volume averaged normalised Q-criteria, (B) depicts ratio of volume averaged normalised Q-criteria with baseline, (C) depicts the maximum instantaneous Q-criteria (in sec^-2^) and (D) depicts ratio of maximum instantaneous Q-criteria with baseline, evaluated at the timepoint corresponding to the peak systole in the respective baseline pulsatile flow (E) Isosurfaces of Normalised Q-criteria > 0.1%Q_max_ (in sec^-2^) for patients with a known stroke or cerebrovascular(CVA) outcome after LVAD deployment(top row) and patients with non stroke/CVA outcome(bottom row).

### 3.4 Comparison of state of wall shear due to LVAD flow

Figure 10 presents the analysis and comparison of wall shear stress along the aorta endothelial lining across the *CVA, non-CVA*, and pre-implantation *baseline* cases. Figure 10.a. presents the averaged wall shear descriptor ⟨Φ_*t*_⟩ as defined in Section 2.4 compared across the 6 *CVA* cases and the 6 *non-CVA* cases. We observe a higher mean value for the averaged wall shear descriptor ⟨Φ_*t*_⟩ for the *CVA* cases (*mean =* 0.17 *g*.*mm*^*-1*^.*sec*^*-2*^) as compared to the *non-CVA* cases (*mean =* 0.14 *g*.*mm*^*-1*^.*sec*^*-2*^). This indicates that aortic hemodynamics driven by an operating LVAD is potentially associated with generally elevated levels of wall shear along the aorta on an average for cases with a stroke outcome when compared to cases without a stroke outcome. The ratio of the averaged wall shear descriptor between the LVAD models and the baseline models are compared in Figure 10.b. Contrary to the absolute values of ⟨Φ_*t*_⟩,we observe that the ratios with baseline are lower in average for the *CVA* cases (*mean =* 1.30) when compared to the *non-CVA* cases (*mean =* 1.54). These observations suggest that cases with a stroke outcome after LVAD implantation had an elevated state of wall shear under the estimated baseline flow conditions, when compared to cases without a stroke outcome. Figure 10.c. presents a 3-dimensional visualization of the TAWSS distribution along the lumen surface of the aortic arch for the *CVA* and *non-CVA* cases. We observe the expected high wall shear values at the LVAD outflow jet impingement location, extremely low wall shear in the regions of stasis as indicated in Figure 6, and the variation in wall shear distribution patterns depending upon the flow development post-impingement which is a function of LVAD outflow graft anastomosis angles and LVAD flow rate. Like Sections 3.2 and 3.3, Table 3 presents a list of descriptive statistics for these quantities for further quantitative reference supporting our observations. Additional details on statistical analysis of the descriptors are included in the appendix to the manuscript.

**Figure 10.**
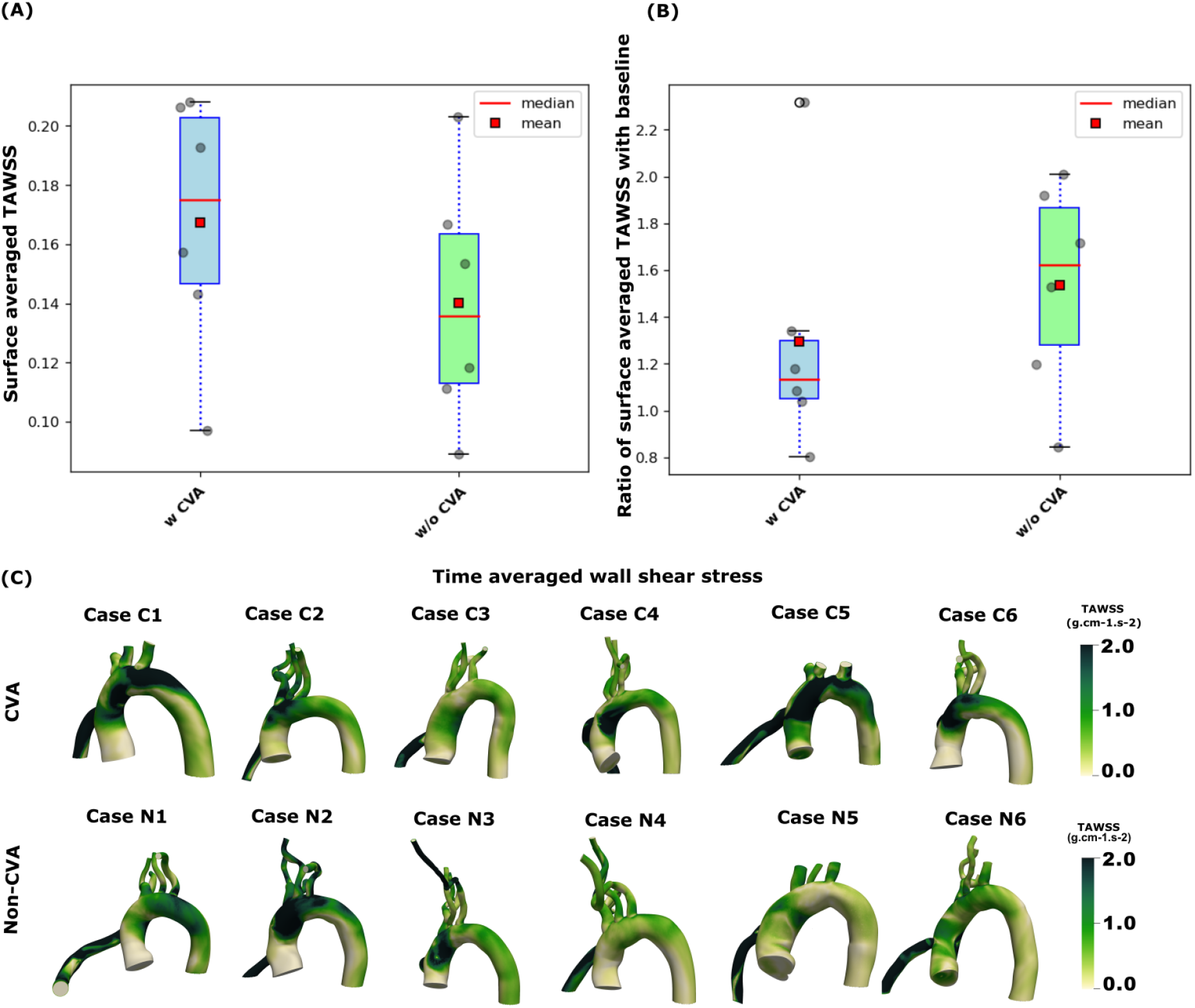
Comparison between trends in wall shear stress (in units of g.mm^-1^.sec^-2^) for CVA vs. non-CVA cases: (A) on the left shows surface and time averaged wall shear stress values with LVAD graft; (B) presents the ratio of surface and time averaged wall shear stress values with that of the baseline models; and (C) illustrates the time averaged wall shear stress (in g.mm^-1^.sec^-2^) visualized across the aortic arch, highlighting elevated WSS in the LVAD outflow jet impingement zones.

### 3.5 Combined analysis of hemodynamic alterations from estimated baseline

Here, we illustrate a comparative analysis of the different hemodynamic descriptors considered for our study by considering their differences in a combined manner, as opposed to looking at them individually. Specifically, we isolated the descriptors that comprise a ratio between LVAD-driven flow variables and corresponding variables for an estimated baseline flow. As we described in our prior work [32], we can create a cumulative quantitative metric as a surrogate for the extent of hemodynamic alteration, by summing up the deviations from unity for each of these descriptor ratios defined as follows:

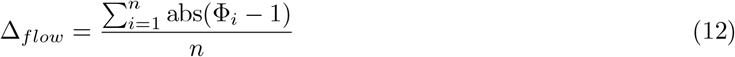

where Φ_*i*_ is a placeholder for the ratios of helicity, vorticity, and wall shear descriptors; and *n* denotes the number of such descriptors being considered for the analysis. If the individual ratios are closer to 1.0, Δ_*flow*_ will end up being a small number, indicating low levels of bulk differences between LVAD-driven and baseline flow. Conversely, larger departures from 1.0, will lead to large Δ_*flow*_ indicating greater bulk difference between LVAD-driven and baseline flow. We computed the above for the ratios of three descriptors: ⟨Φ_*H*_⟩ for positive helicity, ⟨Φ_*Q*_⟩ for vorticity, and ⟨Φ_*t*_⟩ for wall shear; and the resulting Δ_*flow*_ data is presented in Figure 11. We observe that the resulting descriptors are lower for cases with a stroke (*mean = 0*.*75*) as compared to those without a stroke (*mean=1*.*76*). This provides an illustration of how the combined analysis presented in this study can be used to develop a metric or model that can help classify cases with and without stroke outcome for future analysis.

**Figure 11.**
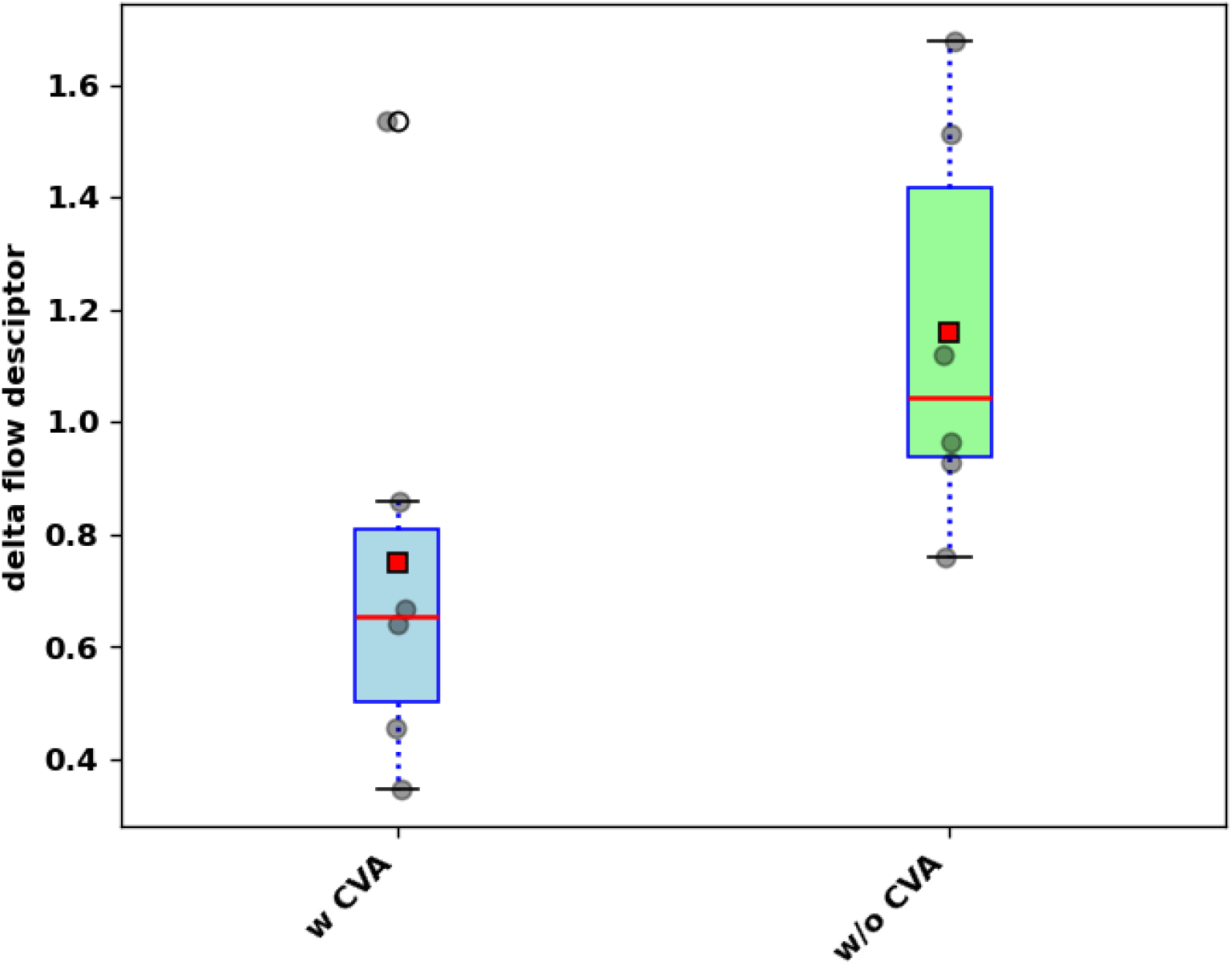
Comparison between the combined descriptor for hemodynamic alterations referred to as Δ_flow_ - a combination of bulk differences between helicity (positive), vorticity, and wall shear stress; computed between the LVAD-driven state of flow, and estimated baseline or pre-LVAD state of flow.

## 4 Discussion

We have presented a systematic retrospective quantitative comparison of aortic hemodynamics between a small cohort of subjects who did and did not report a stroke outcome after LVAD implantation. Our analysis comprised two categories of flow information. First, we quantified hemodynamics in the aorta driven by an operating LVAD for these two groups of subjects. Second, we derived an estimate of what aortic hemodynamics in these subjects would have been without LVAD support (*referred to here as the baseline models*). In prior works, we have interpreted these differences using a conceptual two jet flow model for LVAD hemodynamics [32, 33]. Specifically, flow during LVAD operation is driven by a jet moving into the aorta from the LVAD outflow graft, while under normal ventricle operation, the flow into the aorta is driven by a different jet originating from the systolic opening of the aortic valve. The differences in hemodynamics between these two configurations can be viewed as the hemodynamic alterations experienced by the vascular system of a subject who transitions into LVAD circulatory support. Results from our analysis here show that hemodynamics driven by an operating LVAD, as well as the extent of hemodynamic alterations from an estimated baseline flow, can potentially indicate key differences when compared against stroke outcomes post-LVAD implantation. While we presented evidence of differences in the hemodynamic descriptors between cases with and without stroke, we did not infer broader statistical generalizations owing to the small sample size of 6 patients in each group. This, however, remains a focus of our ongoing investigations using the methodology and analysis developed in this study.

The results also link to a deeper mechanistic interpretation when considering stroke etiology in patients on LVAD support, and other vascular pathological features due to an operating LVAD. Ischemic stroke in patients on LVAD support can originate from thromboemboli that move into the aorta from the LVAD outflow graft (*likely from stagnated blood in the dysfunctional ventricle*), and are subsequently transported to the brain leading to a stroke. Another source of stroke lies in increased clotting propensity within arterial hemodynamics because of the way flow in the aorta is driven by LVAD outflow jet impingement. Increased thrombogenicity is commonly indicated by higher flow stasis, and less mixing of platelets and underlying biochemical species in the flow. Hemodynamic features such as helicity and vortex dynamics can play a role in both mechanisms. From a flow physics perspective, higher vorticity can indicate lower extent of stasis and higher helicity can foster greater extent of mixing. Also, increased vortical and swirling flow can increase the chances of finite size thromboembolic particles to move towards the cervical vessels into the brain. Additionally, elevated levels of wall shear stress can indicate more systemic effects in vascular health, beyond mechanisms associated with thrombogenicity and thromboembolisms. These include triggering inflammatory responses, endothelial trauma and dysfunction, atherosclerotic potential, and even influencing hormonal regulation and endocrine signaling [45–47]. We note that, while our approach here was to directly use grid-based flow data to devise Eulerian descriptors for flow features (*such as flow stasis*), further analysis using Lagrangian particle-based techniques can also be conducted to develop additional quantitaive descriptors (*such as residence time denoting flow stasis*). However, this involves additional computational cost for the integration of a sufficiently large number of particles through the flow field, and hence for the scope of this present study, we did not include such additional variables in our analysis. This is not inherently a limitation of our modeling approach presented here.

The evidence that hemodynamics potentially holds quantifiable hidden information about stroke risks for patients on LVAD circulatory support has broad implications. This provides valuable insights to indicate that factors contributing to stroke risks post-implantation can involve both blood-pump/device interactions, as well as hemodynamic features downstream from the pump. This could inform future advances in the personalization of surgical implantation of LVAD, the assessment of LVAD treatment outcomes, and the monitoring and long-term management of patient health on LVAD support. In particular, hemodynamic descriptors obtained from such *in silico* virtual twin models can be used to inform a stratification model for stroke risks, that can enable surgical decision making and estimates of outcomes. While this was not the focus of this study, we have include in the supplementary data a potential starting point for such analyses for additional reference for the readers, and as an indication of future utility of our methodology. While our analysis was conducted retrospectively for patients who have undergone LVAD implantation, in prior works we have demonstrated that such analysis can be conducted also in form of virtual prospective surgery [32, 33], where several potential graft anastomoses can be evaluated using hemodynamic descriptors as described here. Specifically, there is major interest in surgical optimization of LVAD implantation. Specifications of the anastomosis between the LVAD outflow graft and the aorta has profound impact on hemodynamic characteristics within the aorta, downstream from the interaction of blood with the LVAD pump parts and surfaces. Leveraging the link between these hemodynamic characteristics (*such as the ones described in this study*) and post-operative stroke, can lead to identification of optimal anastomosis to reduce stroke incidence. Methodology and findings reported here can lead to a virtual twin enabled surgical approach individualized for each patient for such optimization. Additionally, the underlying hemodynamic information discussed here can be used to advance current understanding of stroke etiology in patients on LVAD support. Since stroke etiology is known to have a connection with mechanism of formation of clot and clot composition, this can further inform anticoagulation or antiplatelet therapy as well as inform the need for additional imaging and clinical workup. Furthermore, referring to the potential risk stratification model based on the hemodynamic features, post-operative anticoagulation and monitoring can potentially be tailored to the patient’s individual likelihood of stroke after LVAD implantation. For instance, using hemodynamic insights on stroke risks, patients at lowest risk of stroke could potentially be managed with lower levels of anticoagulation, thereby reducing the incidence of bleeding complications. We note that the *in silico* methodology devised here is able to quantitatively extrude the hidden connection between flow and stroke outcomes because of its ability to generate well-resolved space-time varying hemodynamic information post-LVAD implantation. Such detailed flow information is currently unavailable from standard-of-care imaging. 4D flow MRI currently provides the most resolved flow information amongst existing imaging techniques, but most LVADs are currently not MRI compatible. Lastly, the higher ratios of Q-criteria and wall shear stress with baseline in *non-CVA* cases compared to *CVA* cases hints at higher flow vorticity and shear stress in these particular patients under normal aortic inlet condition. Such information enables linking pre-implantation hemodynamic status to post-implantation stroke risks, which is of high significance in terms of surgical planning and treatment efficacy assessment.

Our analysis is based on patient medical images, clinical variables, and outcomes; leading to a systematic personalized digital representation of each individual patient with an operating LVAD. In its classical notion as discussed in several recent perspective reviews [20–22], each digital twin should represent an individual but not population. Here, we seek to advance this notion by presenting an example of virtual composition of a cohort using individual digital twins. Specifically, each of our 12 models are personalized twins, leading to the virtual cohort of 6 patients each, with and without stroke outcomes. Additionally, through the development of the pre-implantation *baseline model* for flow, we demonstrate one innovative approach where the digital twins can be used to conduct markedly unique *‘what if ‘* scenario assessment, that would not be possible with other existing techniques alone. Finally, we note that we have not focused on analysis that leads to a two-way interaction between the digital twin and patient data. This would, for instance, be a suitable case for personalized surgical planning and post-surgical monitoring. Such advancements will comprise complicated, yet valuable, next steps for data-integrated *in silico* modeling investigations such as the study presented here.

Our study was associated with several underlying assumptions and limitations which are noted here. *<u>First:</u>* we were unable to incorporate any effect of aortic insufficiency and aortic valve opening, owing to the difficulty in obtaining such information for patient cases. This remains an ongoing question of interest for us, and is not an inherent limitation of the computational framework as any such evidence of valve opening can be easily integrated into our hemodynamics analysis framework presented here. *<u>Second:</u>* we conducted our study on a relatively small cohort of 12 patients, divided into 6 for each categories (*with/without stroke*). Currently ongoing efforts are aimed at including more clinical cases into our study, and thereafter conducting more comprehensive statistical analysis and hypothesis testing with the larger sample set of data. With the approach as presented here, such analysis on larger patient cohorts can enable development of quantitative metrics and building statistical inference models or neural network surrogate models that pre-surgically relate post-surgical hemodynamics to clinical outcomes. *<u>Third:</u>* our hemodynamics modeling itself was based on standard state-of-the-art assumptions in CFD modeling of large artery hemodynamics. Specifically, they include assumptions of the aorta being rigid walls (effectively assuming that the aorta wall displacements are negligible in determining the state of hemodynamics as described here); and the rheology of blood being linear Newtonian (a widely acknowledged and accepted assumption in large artery hemodynamics modeling, although local elevated shear rates in the jet impingement region may cause fluctuations in rheology which was not accounted for). Additionally, improvements in subject specific estimates of the boundary data such as inlet flow profile at the LVAD outflow graft inlet, or outflow boundary conditions, will substantially enhance the quality of these computational predictions. This remains a challenging endeavor for LVAD-driven flows, however, since flow-based imaging modalities such as 4D MRI or ultrasound are currently not compatible with LVADs and/or are not standard-of-care for LVAD patients. *<u>Fourth:</u>* in terms of local hemodynamic effects we have also not accounted for any turbulence closure models that capture local turbulence in flow. This remains an ongoing area of investigation, owing to the higher computational cost and numerical complexities of accounting for turbulence closure models in vascular applications; as well as including the effects of microscale suspension nature of blood into the turbulence closure description. The underlying Reynolds number here is in the regime of 1000-3000 as also noted in prior works [30], which is in the transitional regime, and not fully turbulent regime. Hence, by sufficient resolution of mesh, we are still able to capture a substantial extent of the underlying non-linearity and vortex structure scales, as also indicated in prior related works [28]. *<u>Finally:</u>* the scope of this work was focused on demonstrating our digital twin cohort based analysis of hemodynamic differences with respect to stroke outcomes. Keeping with this scope, we have not conducted any benchtop or *in vivo* validation for the LVAD models. However, the underlying SimVascular solvers and components have been validated against experimental data in several prior works [48]. Additional validation for LVAD models will continue to motivate our ongoing and future efforts.

## 5 Concluding remarks

In summary, we have presented a computational retrospective hemodynamic analysis for a cohort of patients supported with LVADs, and report on factors associated with postoperative stroke. Our *in silico* analysis integrated patient CT image-based modeling, finite element computational flow modeling, and clinical and outcomes data towards creating digital twins of each patient. We characterized flow post-implantation, as well as obtained an estimate of pre-implantation flow without an LVAD (*referred to as baseline models*). We found that across the models considered, descriptors of positive helical flow, extent of vorticity, and extent of wall shear, all had higher average values for cases with a stroke outcome. When compared against estimated baseline flow pre-implantation, extent of alteration in positive helicity was higher in patients with stroke, while the alteration in vortical flow and wall shear was lower. These observations collectively provide evidence that hemodynamics can reveal quantifiable connections and differences when correlated against stroke outcomes in subjects on LVAD support. Understanding the patient-pump interaction and how blood flow in the aorta is impacted by the presence of the LVAD can identify modifiable risk factors to ultimately improve patient outcomes.

## Supporting information

Supplementary-Animation: Velocities Combined

## Data Availability

All simulations have been conducted using the open source SimVascular software tool. Imaging and quantitative simulation data that support the findings of this study are available from the corresponding author upon reasonable request.

## Ethics Statement

This study used patient data in an anonymized form, solely for retrospective secondary computational analysis. The study was declared to be exempt from additional IRB review by the Colorado Multiple Institutional Review Board (COMIRB).

## Funding Acknowledgements

This work was supported by a University of Colorado Anschutz-Boulder (AB) Nexus Research Collaboration Grant awarded to DM and JP; as well as partially supported by a National Institutes of Health Award (R21EB029736) awarded to DM. This work utilized resources from the University of Colorado Boulder Research Computing Group, which is supported by the National Science Foundation (awards ACI-1532235 and ACI-1532236), the University of Colorado Boulder, and Colorado State University.

## Author Contributions

AS conducted all modeling, simulation, and quantitative analysis for the LVAD models. SM expanded the analysis by including the baseline models. KC supported post-processing of all hemodynamic data from the LVAD models. JP and EM guided study design, patient selection, hemodynamic parameter selection, clinical records interpretation, and clinical interpretation of hemodynamic data. DM designed the study, analysed and interpreted simulation data, and drafted the manuscript in collaboration with AS, SM, KC, EM, and JP. All authors reviewed and finalized the draft, and are in agreement regarding the final content of the manuscript.

## Appendix: Discussion on statistical analysis

Here we provide some additional discussions and commentary on statistical analysis of the hemodynamic descriptors computed for the 12 patient virtual twin cohort. Given the small sample size of 12 patients, decomposed into 6 patients each for stroke vs no-stroke outcomes, a detailed statistical inference analysis or risk stratification modeling was not the focus of this study. However, to demonstrate potential utility of analysis using the kind of data obtained from our model (*as such efforts extend to larger subject cohorts*) we are presenting here some additional details on relevant statistical analysis. Of note, we conducted a check on whether the individual samples are normally distributed - for each hemodynamic descriptor - using a Shapiro-Wilk test. For the small sample size, we rejected the null hypothesis of normality at a p-value of 0.01 or lesser as opposed to 0.05 (*that is, if p-value is less than 0*.*01 from the test, the distribution is strongly not normal*). For those descriptors for which Shapiro-Wilk test rejected normality, we used a non-parametric Mann Whitney U test to determine significance level of the difference between the *CVA* and *non-CVA* subjects. For the other descriptors, we used the Welch’s t-test for determining significance level of the differences. The individual data and p-values from these tests are all included in Figure 1 provided below.

Additionally, for demonstration purposes, we explore the utility of the hemodynamic descriptors to create a model for stroke identification using simple logistic regression. We set the *CVA* vs *non-CVA* cases using a categorical variable *Y* with values of 1 and 0 respectively. Here, for demonstration, we take as the input *X* the descriptor for combined hemodynamic alteration Δ_*flow*_ described in the main manuscript (*refer also to spreadsheet in the suplementary dataset for details*). Using the 12 sample categorical data pair of *Y* vs *X*, we fit a logistic regression model. We then create a random uniform sample of independent *X* for prediction, and generate the categorical prediction of *CVA* vs *non-CVA* using the logistic model. The resulting predictions into a stroke vs no-stroke categorization is presented in Figure 2 below. As we grow such datasets to include more patient samples, we can expand this analysis by assessment of prediction accuracy of this model using a confusion matrix. We wrap this discussion with the note that this discussion of statistical analysis is to supplement the quantitative analysis presented in the main manuscript, and that owing to the small sample sizes considered here, we cannot predict definitive statistical significance with sufficient power (and it was not our objective to conduct a similar detailed statistical analysis for this study).

## Supplementary Material Information

### S1 Data spreadsheet

We have included a labelled spreadsheet - *“Dataset-hemodynamic-descriptors*.*xlsx”* - containing the dataset presented in this manuscript. The workbook contains two sheets. The first sheet contains the case IDs, associated numerical parameters including boundary conditions, and mesh sizes, and the numerical data for the various hemodynamic descriptors as computed and presented in this paper. The format of the database is as presented in Table 2 in the main manuscript. The second sheet contains the detailed quantitative summary statistics for the hemodynamic descriptors listed in our study, as well as details about test fo normality and statistical tests for differences between stroke and no-stroke outcomes. The format of the database is as presented in Table 3 in the main manuscript.

### S2 Animation of velocity profiles for each patient cases with LVAD

Alongwith the manuscript we have included a separate animation file named *“velocity compiled*.*mp4”*, which presents a combined animation of changes in velocity streamtubes with time in over one second of LVAD operation for all 12 patient cases with an attached LVAD graft. This animation provides a temporally varying visualization of the velocity streamtubes, the image for which at *t* = 0.18 sec time instant of the third cycle is presented in Figure 4 of the main manuscript. The animation video is of 14 seconds duration, and considering the LVAD operating cycle as 1 second, the animation is thus rendered 14 times slower than the actual LVAD operational cycle.

### S3 Animation of vorticity for each patient cases with LVAD

Additionally, we have included a separate animation file named *“vorticity compiled*.*mp4”*, which presents a combined animation of changes in vortex structures with time over one second of LVAD operation for all 12 patient cases with an attached LVAD graft. This animation provides a temporally varying visualization of the vortex structures formed as LVAD outflow graft drives a jet flow into the aortic arch, the image for which at *t* = 0.18 sec time instant of the third cycle is presented in Figure 7 of the manuscript. The animation video is of 14 seconds duration, and considering the LVAD operating cycle as 1 second, the animation is thus rendered 14 times slower than the actual LVAD operating cycle.

### S4 Additional data illustrating LVAD outflow graft reconstruction

For reliable digital twin replication of the subject models as described in this study, it is important to have high quality CT images which includes clearly the LVAD outflow graft ans its connection to the aorta, and from which the aortic geometry as well as the LVAD outflow graft can be reliably reconstructed (thereby making the complete model patient specific). Here, we include a table that notes the average diameter of the reconstructed LVAD outflow graft, presented as an average acropss multiple cross-sections. We include the comparison against commonly known graft diameters, to provide additional quantitative proof of the quality of LVAD outflow graft reconstruction. Additionally, we provide a sequence of images in Figure S1, that depicts the precise reconstruction of the LVAD outflow graft and, critically, its attachment to the aorta, directly based off of the high quality CT images.

**Table S1:** Table showing averaged graft diameter reconstructed from images, and their comparison against commonly known graft diameters, for the different cases. Note that the grafts are expected to expand up to 10% during operation due to flow pressure, from their baseline values.

| Case | Model | Reconstructed dia. (mm) | Expected dia. (mm) | Difference (%) |
| --- | --- | --- | --- | --- |
| C1 | HM2 | 15.30 | 14.00 | 9.00 |
| C2 | HVAD | 11.46 | 10.00 | 14.00 |
| C3 | HM2 | 14.21 | 14.00 | 2.00 |
| C4 | HM3 | 14.59 | 14.00 | 4.00 |
| C5 | HVAD | 10.69 | 10.00 | 7.00 |
| C6 | HVAD | 11.62 | 10.00 | 15.00 |
| N1 | HM3 | 16.43 | 14.00 | 16.00 |
| N2 | HVAD | 10.14 | 10.00 | 1.00 |
| N3 | HM3 | 15.77 | 14.00 | 12.00 |
| N4 | HM3 | 15.16 | 14.00 | 8.00 |
| N5 | HVAD | 11.41 | 10.00 | 13.00 |
| N6 | HM3 | 16.17 | 14.00 | 14.00 |

**Figure S1:**
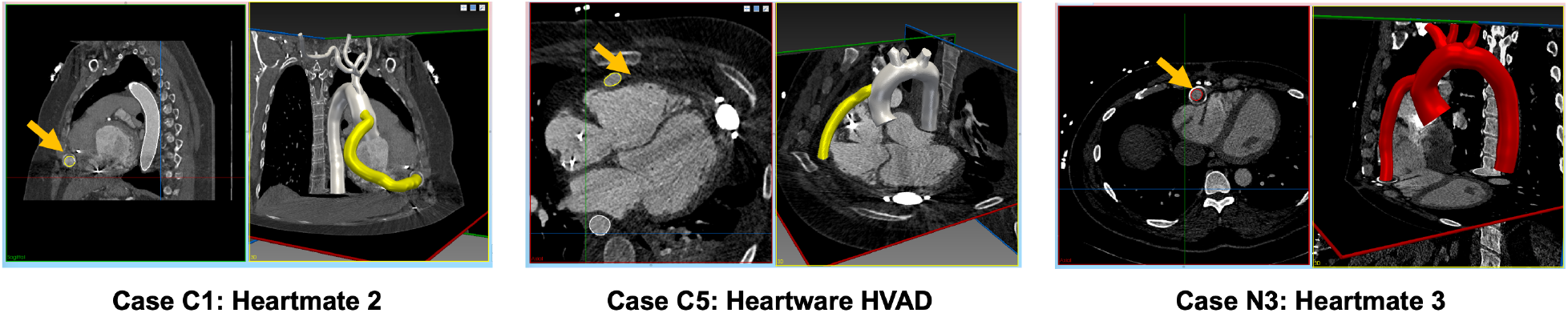
Illustration of image-driven LVAD outflow graft anastomosis reconstruction for digital twin generation. Examples are shown for one each of the three different VAD models used.

### S5 Visualization of hemodynamic descriptors for baseline models

Here we present additional visualizations for hemodynamics data for the 12 baseline models considered in this study. Figure S2 illustrates the simulated hemodynamic patterns for the 12 baseline cases considered in this study, visualized using streamtubes generated at *t* = 0.18 second time point of the third simulation cycle. We observe that for pre-implantation baseline flow conditions driven by jet flow from the aortic root, the aortic hemodynamics have visibly lesser the vortical and helical flow intensity compared to the corresponding LVAD graft implanted cases (Fig. 3) discussed in Section 3.1. This complements the observations reported in Section 3.2 and Section 3.3. Moreover, the velocity in aortic root for baseline cases are higher, and no prominent flow stasis regions in the aortic root are observed, when compared against the corresponding flow for the LVAD implanted counterpart. Figure S3 depicts the isosurfaces of time averaged LNH contours generated at both +0.2 and -0.2 for each of the 12 baseline models. This additional visualization is included to qualitatively compare with the LNH isosurfaces of 12 LVAD implanted models (Fig. 4(C)). Figure S4 represents a 3-dimensional visualization of the time averaged wall shear stress distribution in the aortic arch for the 12 baseline cases. The wall shear stress is observed to be lower and uniformly distributed all over the aortic arch when compared with TAWSS distribution for cases with LVAD implants as shown in Fig. 6(C).

**Figure S2:**
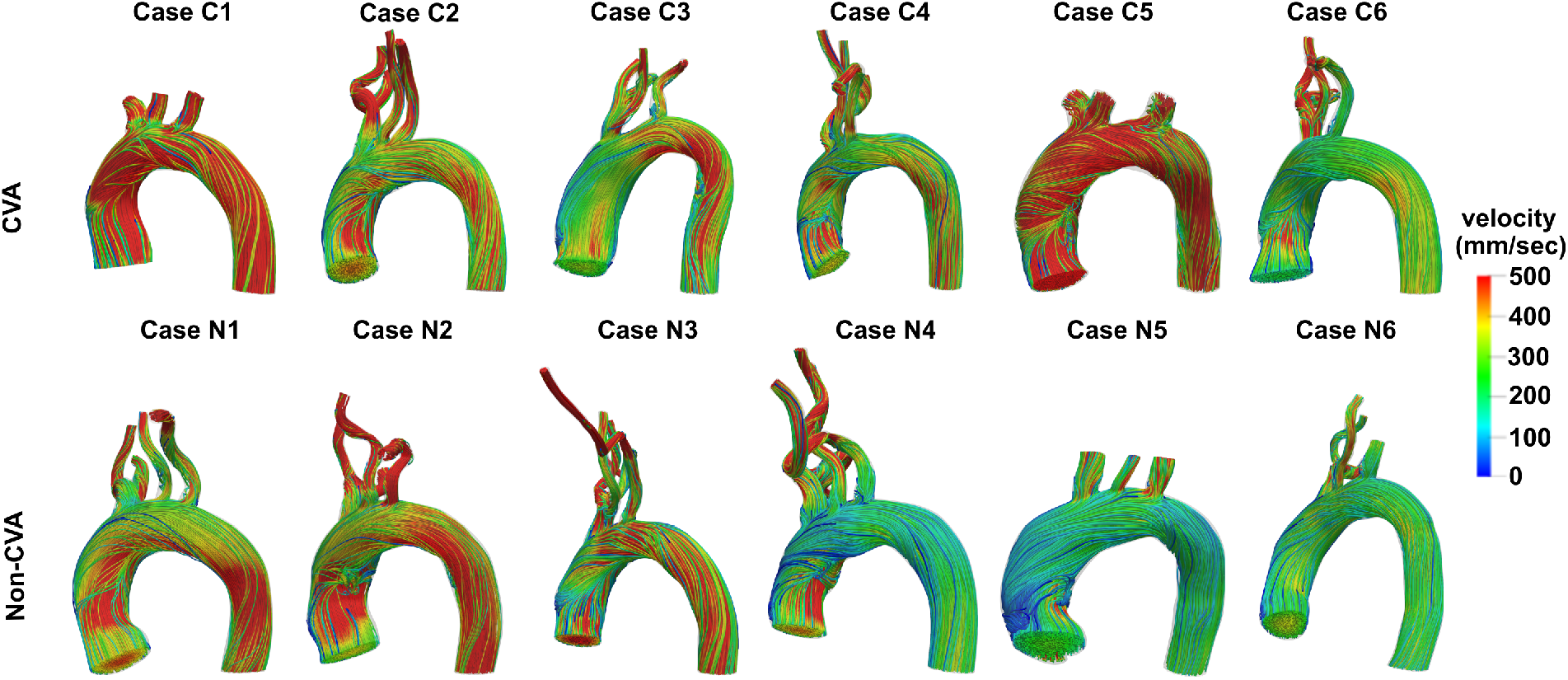
Velocity streamtubes for the estimated baseline flow for the 12 patient cases included in this study.

**Figure S3:**
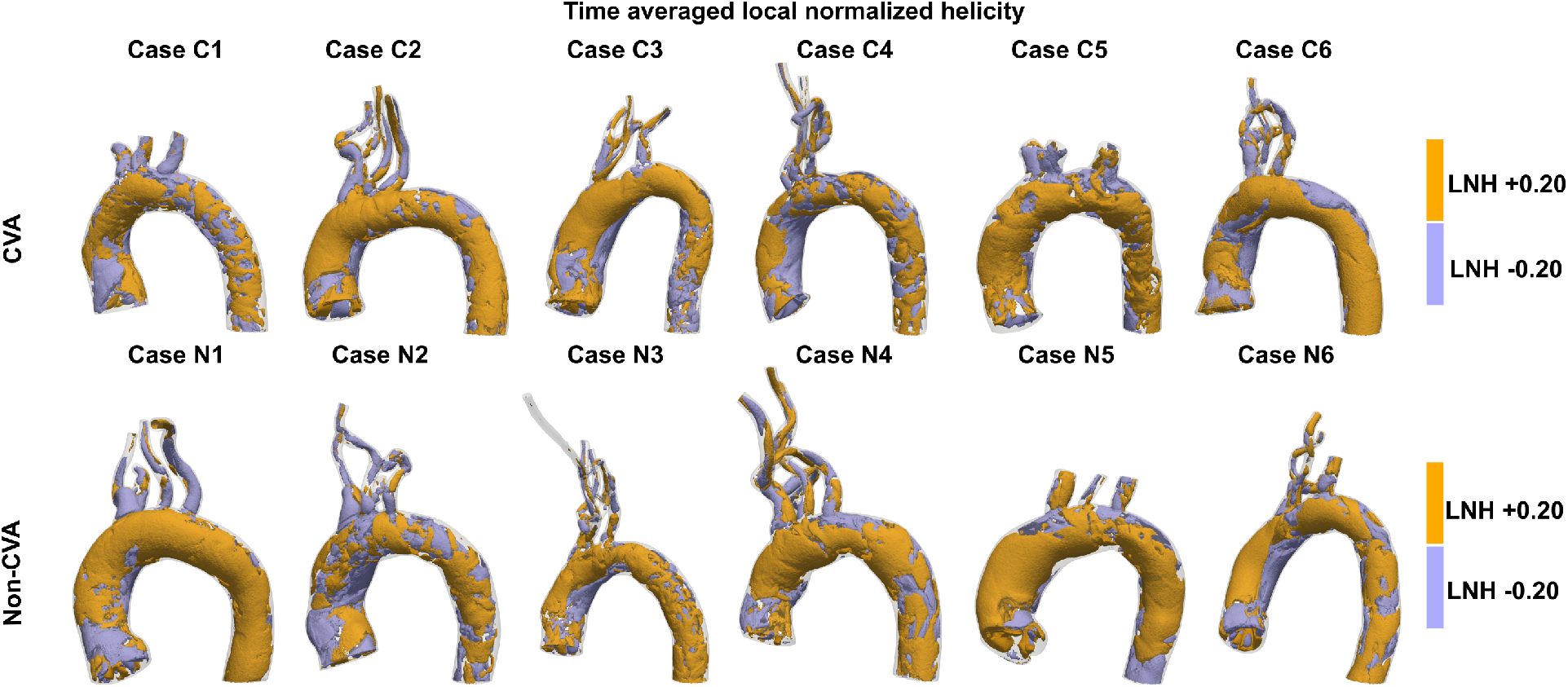
Isosurface visualisations of time averaged local normalised helicity (LNH) for baseline flow for the 12 patient cases included in this study.

**Figure S4:**
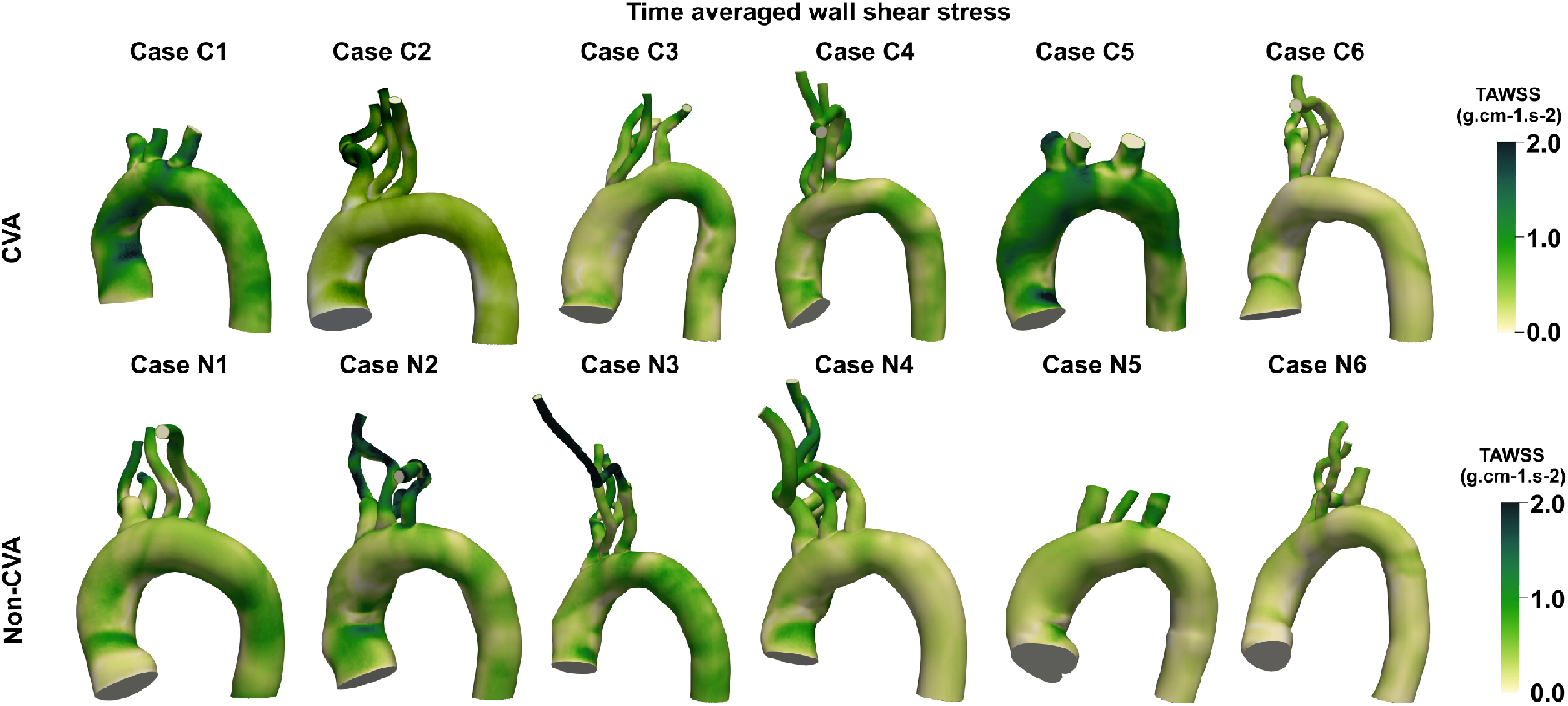
Visualisation of time averaged wall shear stress over the aortic arch for the baseline flow for the 12 patient cases included in this study.

## References

1. Roger, V. L. Epidemiology of heart failure: a contemporary perspective. Circulation Research 128, 1421–1434 (2021).

2. Tsao, C. W., Aday, A. W., Almarzooq, Z. I., Alonso, A., Beaton, A. Z., Bittencourt, M. S., Boehme, A. K., Buxton, A. E., Carson, A. P., Commodore-Mensah, Y., et al. Heart disease and stroke statistics—2022 update: a report from the American Heart Association. Circulation 145, e153–e639 (2022).

3. Rose, E. A., Gelijns, A. C., Moskowitz, A. J., Heitjan, D. F., Stevenson, L. W., Dembitsky, W., Long, J. W., Ascheim, D. D., Tierney, A. R., Levitan, R. G., et al. Long-term use of a left ventricular assist device for end-stage heart failure. New England Journal of Medicine 345, 1435–1443 (2001).

4. Slaughter, M. S., Rogers, J. G., Milano, C. A., Russell, S. D., Conte, J. V., Feldman, D., Sun, B., Tatooles, A. J., Delgado III, R. M., Long, J. W., et al. Advanced heart failure treated with continuous-flow left ventricular assist device. New England Journal of Medicine 361, 2241–2251 (2009).

5. Starling, R. C., Naka, Y., Boyle, A. J., Gonzalez-Stawinski, G., John, R., Jorde, U., Russell, S. D., Conte, J. V., Aaronson, K. D., McGee, E. C., et al. Results of the post-US Food and Drug Administration-approval study with a continuous flow left ventricular assist device as a bridge to heart transplantation: a prospective study using the INTERMACS (Interagency Registry for Mechanically Assisted Circulatory Support). Journal of the American College of Cardiology 57, 1890–1898 (2011).

6. Pagani, F. D., Miller, L. W., Russell, S. D., Aaronson, K. D., John, R., Boyle, A. J., Conte, J. V., Bogaev, R. C., MacGillivray, T. E., Naka, Y., et al. Extended mechanical circulatory support with a continuous-flow rotary left ventricular assist device. Journal of the American College of Cardiology 54, 312–321 (2009).

7. Kormos, R. L., Cowger, J., Pagani, F. D., Teuteberg, J. J., Goldstein, D. J., Jacobs, J. P., Higgins, R. S., Stevenson, L. W., Stehlik, J., Atluri, P., et al. The Society of Thoracic Surgeons Intermacs database annual report: evolving indications, outcomes, and scientific partnerships. The Journal of Heart and Lung Transplantation 38, 114–126 (2019).

8. Kirklin, J. K., Naftel, D. C., Pagani, F. D., Kormos, R. L., Stevenson, L. W., Blume, E. D., Myers, S. L., Miller, M. A., Baldwin, J. T. & Young, J. B. Seventh INTERMACS annual report: 15,000 patients and counting. The Journal of Heart and Lung Transplantation 34, 1495–1504 (2015).

9. Frontera, J. A., Starling, R., Cho, S.-M., Nowacki, A. S., Uchino, K., Hussain, M. S., Mountis, M. & Moazami, N. Risk factors, mortality, and timing of ischemic and hemorrhagic stroke with left ventricular assist devices. The Journal of Heart and Lung Transplantation 36, 673–683 (2017).

10. Harvey, L., Holley, C., Roy, S. S., Eckman, P., Cogswell, R., Liao, K. & John, R. Stroke after left ventricular assist device implantation: outcomes in the continuous-flow era. The Annals of Thoracic Surgery 100, 535–541 (2015).

11. Willey, J. Z., Gavalas, M. V., Trinh, P. N., Yuzefpolskaya, M., Garan, A. R., Levin, A. P., Takeda, K., Takayama, H., Fried, J., Naka, Y., et al. Outcomes after stroke complicating left ventricular assist device. The Journal of Heart and Lung Transplantation 35, 1003–1009 (2016).

12. Acharya, D., Loyaga-Rendon, R., Morgan, C. J., Sands, K. A., Pamboukian, S. V., Rajapreyar, I., Holman, W. L., Kirklin, J. K. & Tallaj, J. A. INTERMACS analysis of stroke during support with continuous-flow left ventricular assist devices: risk factors and outcomes. JACC: Heart Failure 5, 703– 711 (2017).

13. Morgan, J. A., Brewer, R. J., Nemeh, H. W., Gerlach, B., Lanfear, D. E., Williams, C. T. & Paone, G. Stroke while on long-term left ventricular assist device support: incidence, outcome, and predictors. ASAIO Journal 60, 284–289 (2014).

14. Mehra, M. R., Goldstein, D. J., Cleveland, J. C., Cowger, J. A., Hall, S., Salerno, C. T., Naka, Y., Horstmanshof, D., Chuang, J., Wang, A., et al. Five-year outcomes in patients with fully magnetically levitated vs axial-flow left ventricular assist devices in the MOMENTUM 3 randomized trial. JAMA 328, 1233–1242 (2022).

15. Mehra, M. R., Uriel, N., Naka, Y., Cleveland Jr, J. C., Yuzefpolskaya, M., Salerno, C. T., Walsh, M. N., Milano, C. A., Patel, C. B., Hutchins, S. W., et al. A fully magnetically levitated left ventricular assist device. New England Journal of Medicine 380, 1618–1627 (2019).

16. Maltais, S., Kilic, A., Nathan, S., Keebler, M., Emani, S., Ransom, J., Katz, J. N., Sheridan, B., Brieke, A., Egnaczyk, G., et al. PREVENtion of HeartMate II pump thrombosis through clinical management: the PREVENT multi-center study. The Journal of Heart and Lung Transplantation 36, 1–12 (2017).

17. Weisel, J. & Litvinov, R. The biochemical and physical process of fibrinolysis and effects of clot structure and stability on the lysis rate. Cardiovascular & Hematological Agents in Medicinal Chemistry (Formerly Current Medicinal Chemistry-Cardiovascular & Hematological Agents) 6, 161–180 (2008).

18. Bartoli, C. R., Giridharan, G. A., Litwak, K. N., Sobieski, M., Prabhu, S. D., Slaughter, M. S. & Koenig, S. C. Hemodynamic responses to continuous versus pulsatile mechanical unloading of the failing left ventricle. ASAIO Journal 56, 410–416 (2010).

19. Soucy, K. G., Koenig, S. C., Giridharan, G. A., Sobieski, M. A. & Slaughter, M. S. Defining pulsatility during continuous-flow ventricular assist device support. The Journal of Heart and Lung Transplantation 32, 581–587 (2013).

20. Coorey, G., Figtree, G. A., Fletcher, D. F., Snelson, V. J., Vernon, S. T., Winlaw, D., Grieve, S. M., McEwan, A., Yang, J. Y. H., Qian, P., et al. The health digital twin to tackle cardiovascular disease—a review of an emerging interdisciplinary field. NPJ digital medicine 5, 126 (2022).

21. Viceconti, M., De Vos, M., Mellone, S. & Geris, L. Position paper From the digital twins in healthcare to the Virtual Human Twin: a moon-shot project for digital health research. IEEE Journal of Biomedical and Health Informatics (2023).

22. Katsoulakis, E., Wang, Q., Wu, H., Shahriyari, L., Fletcher, R., Liu, J., Achenie, L., Liu, H., Jackson, P., Xiao, Y., et al. Digital twins for health: a scoping review. NPJ Digital Medicine 7, 77 (2024).

23. May-Newman, K., Hillen, B., Sironda, C. & Dembitsky, W. Effect of LVAD outflow conduit insertion angle on flow through the native aorta. Journal of Medical Engineering & Technology 28, 105–109 (2004).

24. Inci, G. & Sorgüven, E. Effect of LVAD outlet graft anastomosis angle on the aortic valve, wall, and flow. ASAIO Journal 58, 373–381 (2012).

25. Karmonik, C., Partovi, S., Loebe, M., Schmack, B., Weymann, A., Lumsden, A. B., Karck, M. & Ruhparwar, A. Computational fluid dynamics in patients with continuous-flow left ventricular assist device support show hemodynamic alterations in the ascending aorta. The Journal of Thoracic and Cardiovascular Surgery 147, 1326–1333 (2014).

26. Callington, A., Long, Q., Mohite, P., Simon, A. & Mittal, T. K. Computational fluid dynamic study of hemodynamic effects on aortic root blood flow of systematically varied left ventricular assist device graft anastomosis design. The Journal of Thoracic and Cardiovascular Surgery 150, 696–704 (2015).

27. Aliseda, A., Chivukula, V. K., Mcgah, P., Prisco, A. R., Beckman, J. A., Garcia, G. J., Mokadam, N. A. & Mahr, C. LVAD outflow graft angle and thrombosis risk. ASAIO Journal 63, 14 (2017).

28. Chivukula, V. K., Beckman, J. A., Prisco, A. R., Dardas, T., Lin, S., Smith, J. W., Mokadam, N. A., Aliseda, A. & Mahr, C. Left ventricular assist device inflow cannula angle and thrombosis risk. Circulation: Heart Failure 11, e004325 (2018).

29. Prather, R., Divo, E., Kassab, A. & DeCampli, W. Computational fluid dynamics study of cerebral thromboembolism risk in ventricular assist device patients: Effects of pulsatility and thrombus origin. Journal of Biomechanical Engineering 143 (2021).

30. Caruso, M. V., Gramigna, V., Rossi, M., Serraino, G. F., Renzulli, A. & Fragomeni, G. A computational fluid dynamics comparison between different outflow graft anastomosis locations of Left Ventricular Assist Device (LVAD) in a patient-specific aortic model. International Journal for Numerical Methods in Biomedical Engineering 31, e02700 (2015).

31. Mahr, C., Chivukula, V., McGah, P., Prisco, A. R., Beckman, J. A., Mokadam, N. A. & Aliseda, A. Intermittent aortic valve opening and risk of thrombosis in VAD patients. ASAIO Journal 63, 425 (2017).

32. Sahni, A., McIntyre, E. E., Pal, J. D. & Mukherjee, D. Quantitative assessment of aortic hemodynamics for varying left ventricular assist device outflow graft angles and flow pulsation. Annals of Biomedical Engineering 51, 1–18 (2023).

33. Sahni, A., McIntyre, E. E., Cao, K., Pal, J. D. & Mukherjee, D. The Relation Between Viscous Energy Dissipation And Pulsation For Aortic Hemodynamics Driven By A Left Ventricular Assist Device. Cardiovascular Engineering and Technology, 1–17 (2023).

34. Updegrove, A., Wilson, N., Merkow, J., Lan, H., Marsden, A. & Shadden, S. Simvascular: An Open Source Pipeline for Cardiovascular Simulation. Annals of Biomedical Engineering 45, 525–541 (2017).

35. Les, A. S., Shadden, S. C., Figueroa, C. A., Park, J. M., Tedesco, M. M., Herfkens, R. J., Dalman, R. L. & Taylor, C. A. Quantification of hemodynamics in abdominal aortic aneurysms during rest and exercise using magnetic resonance imaging and computational fluid dynamics. Annals of Biomedical Engineering 38, 1288–1313 (2010).

36. Mukherjee, D., Jani, N. D., Narvid, J. & Shadden, S. C. The role of circle of Willis anatomy variations in cardio-embolic stroke: a patient-specific simulation based study. Annals of Biomedical Engineering 46, 1128–1145 (2018).

37. Ku, D. N. et al. Blood flow in arteries. Annual Review of Fluid Mechanics 29, 399–434 (1997).

38. Brooks, A. N. & Hughes, T. J. Streamline upwind/Petrov-Galerkin formulations for convection dominated flows with particular emphasis on the incompressible Navier-Stokes equations. Computer Methods in Applied Mechanics and Engineering 32, 199–259 (1982).

39. Franca, L. P., Frey, S. L. & Hughes, T. J. Stabilized finite element methods: I. Application to the advective-diffusive model. Computer Methods in Applied Mechanics and Engineering 95, 253–276 (1992).

40. Franca, L. P. & Frey, S. L. Stabilized finite element methods: II. The incompressible Navier-Stokes equations. Computer Methods in Applied Mechanics and Engineering 99, 209–233 (1992).

41. Taylor, C. A., Hughes, T. J. & Zarins, C. K. Finite element modeling of blood flow in arteries. Computer Methods in Applied Mechanics and Engineering 158, 155–196 (1998).

42. Zamir, M. in The physics of pulsatile flow 67–112 (Springer, 2000).

43. Gallo, D., Steinman, D. A., Bijari, P. B. & Morbiducci, U. Helical flow in carotid bifurcation as surrogate marker of exposure to disturbed shear. Journal of Biomechanics 45, 2398–2404 (2012).

44. Jeong, J. & Hussain, F. On the identification of a vortex. Journal of Fluid Mechanics 285, 69–94 (1995).

45. Hasin, T., Matsuzawa, Y., Guddeti, R. R., Aoki, T., Kwon, T.-G., Schettle, S., Lennon, R. J., Chokka, R. G., Lerman, A. & Kushwaha, S. S. Attenuation in peripheral endothelial function after continuous flow left ventricular assist device therapy is associated with cardiovascular adverse events. Circulation Journal 79, 770–777 (2015).

46. Uzarski, J. S., Scott, E. W. & McFetridge, P. S. Adaptation of endothelial cells to physiologically-modeled, variable shear stress. PloS One 8, e57004 (2013).

47. Millon, A., Sigovan, M., Boussel, L., Mathevet, J.-L., Louzier, V., Paquet, C., Geloen, A., Provost, N., Majd, Z., Patsouris, D., et al. Low WSS induces intimal thickening, while large WSS variation and inflammation induce medial thinning, in an animal model of atherosclerosis. PloS One 10, e0141880 (2015).

48. Lan, I. S., Liu, J., Yang, W., Zimmermann, J., Ennis, D. B. & Marsden, A. L. Validation of the reduced unified continuum formulation against in vitro 4D-flow MRI. Annals of Biomedical Engineering 51, 377–393 (2023).

